# Comparison of Two Genome-Wide Association Studies for Heart Rate Response to Exercise from the UK Biobank

**DOI:** 10.1101/2021.07.07.21259806

**Authors:** Abhinav Thakral, Andrew D. Paterson

## Abstract

The short-term changes in heart rate (HR) during and after exercise are important physiologic traits mediated via the autonomic nervous system. Variations in these traits are associated with mortality from cardiovascular causes. We conducted a systematic review of genome-wide association studies for these traits (with >10,000 participants) with the aim of comparing Polygenic Risk Scores (PRS) from different studies. Additionally, we applied the **ST**rengthening of **R**eporting of **G**enetic **A**ssociation Studies (STREGA) statement for assessing the completeness of reporting of evidence. Our systematic search yielded two studies (Verweij et al. and Ramirez et al.) that met our inclusion criteria. Both were conducted on the UK Biobank. Both defined their exercise traits as the difference between resting HR and the maximum HR during exercise. Their recovery traits were defined differently. Verweij et al. defined 5 recovery traits as the differences between the peak HR during exercise and the HRs at 10-50 sec post exercise cessation. Ramirez et al. defined their recovery trait as the difference between peak HR during exercise and the minimum HR during the minute post exercise cessation. While Ramirez et al. divided their sample into discovery and replication subsets, Verweij et al. analyzed the whole sample together. In terms of results, there were several common SNPs identified between studies and traits. There was evidence for the phenomenon of winner’s curse operating for a SNP from the Ramirez study’s HR recovery analysis. Many of the SNPs were mutually exclusive between the studies. However, there was a good agreement of PRS from the studies. The differences in the results could be attributed to the different exclusion criteria, analytic approaches, and definitions of traits used. Both studies had an under-representation of individuals of non-European ancestry compared to those of European ancestry. Further studies with proportionate representation of individuals of all ancestries would help address this gap.

**Graphical Abstract:** 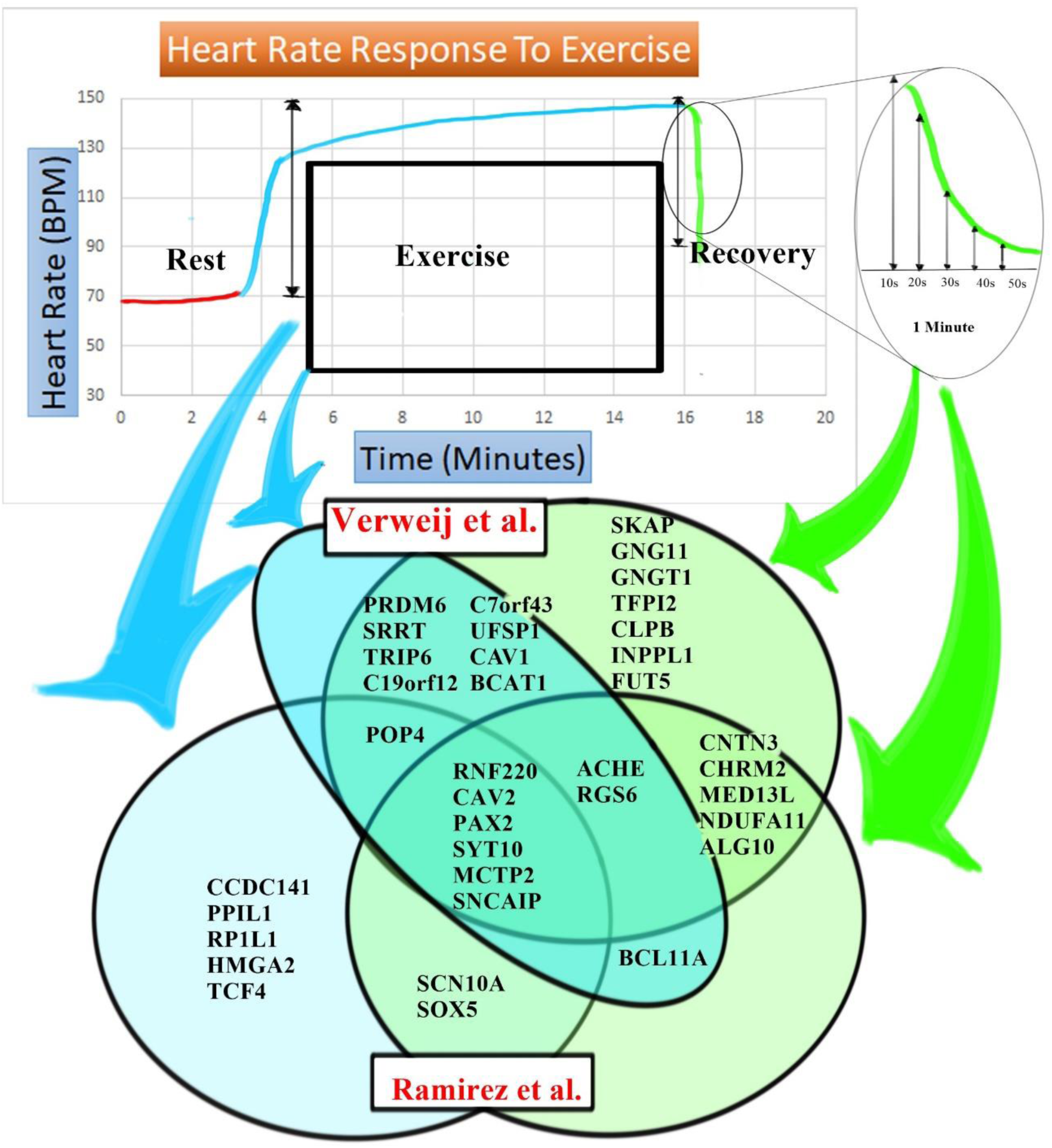

## 1. Introduction

The short-term changes in heart rate (HR) during and after exercise are important physiologic traits. These responses are mediated via the autonomic nervous system(1–3). Variations in these responses are associated with mortality from cardiovascular causes (4–9). Further, these responses have been shown to be heritable using twin (10, 11), and family (12, 13) studies, and have been studied using genome-wide association studies (GWAS)(14, 15).

GWAS are one of the most powerful tools to identify genetic variants associated with phenotypes. Two recently published GWAS (14, 15) that analyzed data from the UK Biobank (16) have identified variants associated with HR responses during and after exercise, some of which are near genes involved in the autonomic nervous system. Both studies also reported polygenic risk scores (PRS) associated with these traits.

Polygenic models for risk prediction derived from GWAS have been shown to predict risk for several common diseases (17). Thus, polygenic scores that predict HR response during and after exercise could be used to assess related traits and cardiovascular health. In general, GWAS with large sample sizes are necessary to generate PRS with a good prediction(17).

We therefore systematically reviewed GWAS of short-term HR response during and after exercise to develop PRS for these traits based on large datasets (>10,000 participants).

The second goal of our review was to compare the PRS from individual GWAS. We wanted to compare the PRS from different GWAS because replication is an essential component of the scientific process. One goal of replication is to confirm or refute the statistical evidence for association (18). However, for several reasons, the findings from the discovery sample may fail to reproduce in a replication sample. One such reason is the “winner’s curse” (19). This phenomenon refers to the lack of reproducibility because the estimates of effects at loci are biased upwards with increasing statistical significance thresholds (20). This bias is most extreme when sample size is small, minor allele frequency (MAF) is low, the significance criterion is stringent, and the magnitude of true genetic association is low (21). Therefore, subsequent studies may be underpowered to detect the association because their power is based on sample size calculations that use the upwardly biased effect estimates from the discovery analysis. Thus, even a true association may fail to replicate in subsequent studies. Therefore, another goal for replication is to improve the estimate of the SNP effect(s) (18).

The final goal of the review was to assess the completeness of reporting of the identified GWAS.

## 2. Methods

We systematically reviewed EMBASE and MEDLINE databases for GWAS examining HR response to exercise and recovery on May 15th, 2020. Appendix 1 provides our search strategy. We restricted our search to studies with a sample size of more than 10,000. We chose a stringent sample size criterion because large sample sizes are required for the generation of PRS with a good prediction. Additionally, large sample sizes are required for minimizing the bias due to the winner’s curse.

### Inclusion Criteria

- Genome-wide Association Studies
- Studies examining HR response to exercise or recovery or both
- Sample Size >10, 000.
- Studies looking at short term changes in HR during exercise

### Exclusion Criteria

- Candidate Gene studies
- Twin and family studies
- Studies not using the GWAS approach
- Studies examining the genetics of resting HR
- Sample size<10,000
- Studies looking at long-term changes in HR in response to exercise

Supplementary Figure 1 provides the flow diagram of our systematic search.

We assessed the completeness of reporting using the guidelines from the STrengthening of Reporting for Genetic Association Studies (STREGA) statement (22),because it is based on existing guidance for reporting observational studies (STROBE) (23). Further, the STREGA statement details the rationale behind decisions and is accessible to scientists from diverse disciplines (22).

We further compared the results of the studies found in the systematic search. We tabulated the SNPs located in the loci common to the studies and compared their P values, beta estimates, and standard errors. Further, we compared the D’ and r^2^ parameters of the SNPs in these loci using LDpair tool from LDLINK (24) using the GBR population from 1000 genomes project phase 3 (version 5) (25)

Finally, we compared the PRS from individual GWAS by looking at their distributions in the European super-population from the 1000 Genomes project. We also performed a Bland Altman analysis (26) to assess agreement between the two PRS.

## 3. Results

Our systematic literature search yielded only two studies (14, 15) that met our inclusion criteria. The included studies by Verweij et al. and Ramirez et al. were conducted on the same population of the UK Biobank (16). Because the two studies examined subsets of subjects from the same Biobank, we provide a comparison of their methods and results.

A comparison of methods used in the studies is summarized in Table 1, which compares their methodology and reporting based on the guidance from the STREGA statement (22). Briefly, both studies were conducted on a subset of the UK Biobank population that participated in the cardio assessment of the UK Biobank. The UK Biobank had 500,000 participants, of which 100,000 were selected for exercise testing, as per protocol (27). Both studies utilized the same imputed genotyping data from the UK Biobank, had similar quality control protocols, and excluded people with conditions or medications that affected heart rate. However, the subsets of participants differed between the studies on account of different exclusion criteria. While Ramirez et al. excluded people with abnormal heart rates (see Table 1), Verweij et al. excluded people who could not exercise beyond a 30-50% level. Further, Ramirez et al. limited their analysis to individuals of European ancestry, while Verweij et al. included everyone, regardless of ancestry, and included 30 principal components as covariates.

**Table 1.** Comparison of methodologies used in the two studies included in the review.

|  | *Ramirez et al.* | *Verweij et al.* |
| --- | --- | --- |
| <u>Setting and Participants:</u> | UK Biobank:<br>a prospective study of 500,000 volunteers, comprising relatively even numbers of men and women aged 40–69 years old at recruitment in the UK (2006-2010). Data collection on phenotypes is ongoing. |  |
| Genotyping | Genotyping was performed by UKB using the Applied Biosystems UK BiLEVE Axiom Array or the UKB Axiom™ Array<sup>41</sup>.<br>DNA was extracted from stored blood samples that had been collected from participants on their visit to a UK Biobank assessment center.<br>Genotyping was carried out by Affymetrix Research Services Laboratory in 106 sequential batches of approximately 4,700 samples.<br>Affymetrix applied a custom genotype calling pipeline and quality filtering optimized for biobank scale genotyping experiments and the novel genotyping arrays, which contain markers that had not been previously typed using Affymetrix technology. This resulted in a set of genotype calls for 489,212 samples at 812,428 unique markers (biallelic SNPs and indels) from both arrays, with which a further quality control and analysis was conducted<br>SNPs were imputed centrally by UKB using the Haplotype Reference Consortium (HRC) panel |  |
| Quality Control | Individuals with high missingness (0.97%), high heterozygosity, and discordance with reported sex and genotyped sex were removed from the analysis. |  |
| Exclusions | Excluded people with abnormal baseline (<40 or >120 bpm), peak (>200 bpm), 1 min post exercise (<40 or >200 bpm) heart rates | Excluded people who could not cycle at 30-50% level (Included people with no risk or minimal risk) |
|  | Excluded people with conditions or medications that could affect heart rate |  |
| Sample Size | ~67k | ~58k |
|  | Sample divided into ~40k discovery and ~27k replication. They also performed joint analysis at top SNPs |  |
| Analysis |  |  |
| Traits | $\Delta HR_{\text{ex}} = \text{PeakHR} - \text{RestingHR}$<br>$\Delta HR_{\text{rec}} = \text{PeakHR} - \text{RecoveryHR}$<br>Resting HR was defined as the mean HR during pre-test period, peak HR as the maximum HR during exercise, and recovery HR as the minimum HR 1 min after the peak exercise. | HR increase = peak HR during exercise - resting HR.<br>HR recovery = maximum HR during exercise - mean HR at $10 \pm 3$, $20 \pm 3$, $30 \pm 3$, $40 \pm 3$, and $50 \pm 3$ s after exercise cessation (HRR10-HRR50).<br>mean resting HR,<br>standard deviation of RR intervals (SDNN, log2 transformed), and<br>root mean square of successive differences between RR intervals (RMSSD, log2 transformed)<br>HR phenotypes were rank-based inverse normal transformed to increase the power to detect low-frequency variants and allow for comparisons of beta coefficients between traits. |
| Covariates | Sex, age, body mass index (BMI), genotyping chip |  |
|  | Resting HR | sex-age interaction, BMI*BMI, the first 30 principal components, the model also included exercise duration, exercise program (30% or 50% maximum |
|  |  | load), maximum workload achieved, and the interaction between exercise program and maximum workload achieved |
| Software Used | BOLT software | BOLT software, R v3.3.2 and STATA/SE Release 13 |
| HWE | $P\text{-value} > 1 \times 10^{-6}$ | -- |
| Imputation | Imputation quality (INFO) > 0.3 |  |
| Missingness | Additional threshold for missingness < 0.0015 | UK BioBank Missingness thresholds used. |
| Population Stratification and Relatedness | Limited Analysis to individuals of European Ancestry (85K)<br>BOLT software accounted for relatedness. In addition, they excluded first and second-degree relatives (kinship coefficient > 0.88) from the replication cohort | Included individuals irrespective of ancestry<br>A conjugate gradient-based iterative framework for fast mixed-model computations was employed to accurately account for population structure and relatedness.<br>Used cluster-robust standard errors with genetic family IDs as clusters.<br>A family ID was given to individuals belonging together based on 3<sup>rd</sup>-degree or closer as indicated by the kinship matrix, which was provided by UK Biobank (kinship coefficient > 0.0442). |
| Threshold for MAF | > 1% |  |
| Model | Additive effects model was assumed |  |
| Significance Thresholds | For Discovery analysis: $P < 1 \times 10^{-6}$ for both traits<br>Replication: $P$ (one-tailed) $\leq 0.05/14 = 2.60 \times 10^{-3}$ for $\Delta\text{HReX}$ and | A conservative genome-wide significant threshold of $p < 8.3 \times 10^{-9}$ was adopted to account for six independent traits |
|  | $\leq 0.05/16 = 3.10 \times 10^{-3}$ for $\Delta HR_{rec}$ and the effect ($\beta$) was in the direction observed in discovery analyses for each trait in the replication cohort.<br>Additional SNPs reported on full dataset analysis at a genome wide significance threshold of $P \leq 5 \times 10^{-8}$. |  |
| Locus Definition | SNPs were mapped to individual loci based on genomic distance of >500 Kb to each side of another SNP.<br>If multiple SNPs fit, only the SNP with the smallest P value was considered for follow-up. | A locus was defined as the highest associated independent SNP ± 1MB |

For the analysis, Ramirez et al. divided the dataset into discovery and replication subsets. They selected the SNPs from the discovery subset based on a Hardy Weinberg equilibrium P>10^-6^, missingness < 0.0015, minor allele frequency (MAF) >0.01, and P ≤ 10^-6^. Next, they tested the significant SNPs from the discovery in the replication using one-tailed tests and corrected for the number of tests. Finally, they analyzed the entire dataset at a genome-wide significance threshold of P ≤ 5 × 10^−8^. On the other hand, Verweij et al. analyzed the entire dataset at a genome-wide significance threshold of p< 8.3 × 10^−9^, assuming independence of all traits.

The outcomes were defined differently across the two traits. For HR response to exercise, Ramirez et al. and Verweij et al. defined their traits identically: HR difference between peak HR during exercise and mean resting HR. While for recovery from exercise, Ramirez et al. defined this as the difference between peak HR during exercise and minimum HR in the minute after exercise cessation. In contrast, Verweij et al. defined five different HR recovery traits as the differences between the peak HR and that at 10s, 20s, 30s, 40s, and 50s post-exercise cessation. Further, they inverse-normal transformed the traits before analysis.

Another important reason for the differences in the subsets of populations analyzed by the two groups was the different criteria used for excluding abnormal or erroneous records of HR responses. Ramirez et al. validated the HR measurements against traits derived from ECG recordings using the algorithm described by Orini et al. (28). For cases with discrepancies between the UK Biobank measurements and the algorithm, they visually inspected ECG recordings. If the QRS complexes were non-identifiable, they rejected the UK Biobank measurements. If the algorithm failed, they corrected the measurements by hand. Ramirez et al. therefore removed 1,482 individuals on account of this mismatch. In contrast, Verweij et al. excluded ECG records without full disclosure data, outlier records with abnormal RR intervals identified based on international recommendations (29), and those identified using an automatic detection method (30). Verweij et al. excluded 2,804 individuals based on abnormal or erroneous HR recordings.

Both studies accounted for sex, age, body mass index, and genotyping chip as covariates in their analyses. However, the analyses in the two studies included several mutually exclusive covariates. Most importantly, while Verweij et al. adjusted for variables related to exercise load and Principal Components, Ramirez et al. adjusted for resting heart rate as a covariate in their analyses.

Ramirez et al. discovered 14 SNPs associated with HR response to exercise and 16 SNPs associated with HR response to recovery in their discovery analysis. Six out of the 14 SNPs replicated for HR response to exercise and seven out of 16 for recovery. In their full analysis, they discovered an additional 8 SNPs associated with the HR response to exercise and an additional 9 SNPs associated with HR response to recovery. Out of a total of 30 SNPs discovered, eight were common to the two traits. In Supplementary Figure 2, we compare the effects (and SE) of the SNPs for the HR response to exercise and recovery in the discovery and replication subsets. The beta estimates of the regression coefficients for both the exercise and recovery traits were not statistically significantly different (P=0.94 and P=0.37, respectively). This result suggests that overall, there was no significant evidence for the winner’s curse in the analysis by Ramirez et al. However, a visual inspection of Supplementary Figure 2A reveals that the SNP in RGS6 (name it) is a distinct outlier. After removing this SNP from the regression analysis (Supplementary Figure 2C), the slope of the regression line increases. The confidence intervals after removing this SNP do not overlap with those before removing the SNP. These findings suggest that the winner’s curse is operating for this SNP. Further, this SNP had the lowest MAF (0.013) amongst those found to be associated with the recovery traits in the analysis by Ramirez et al. The association for this SNP was just below the significance threshold (P= 4.3 ×10^-8^ in the full analysis). These findings are in line with Poirier et al.’s (21) observations on the winner’s curse.

Verweij et al. discovered 23 primary SNPs associated with HR increase or recovery traits. In Figure 1, we compare the effects (and SE) of the primary SNPs at loci common to the two studies for both exercise and recovery traits. Eleven primary SNPs were shared between the studies. Of these, three were for exercise traits, and eight were for recovery traits.

**Figure 1.**
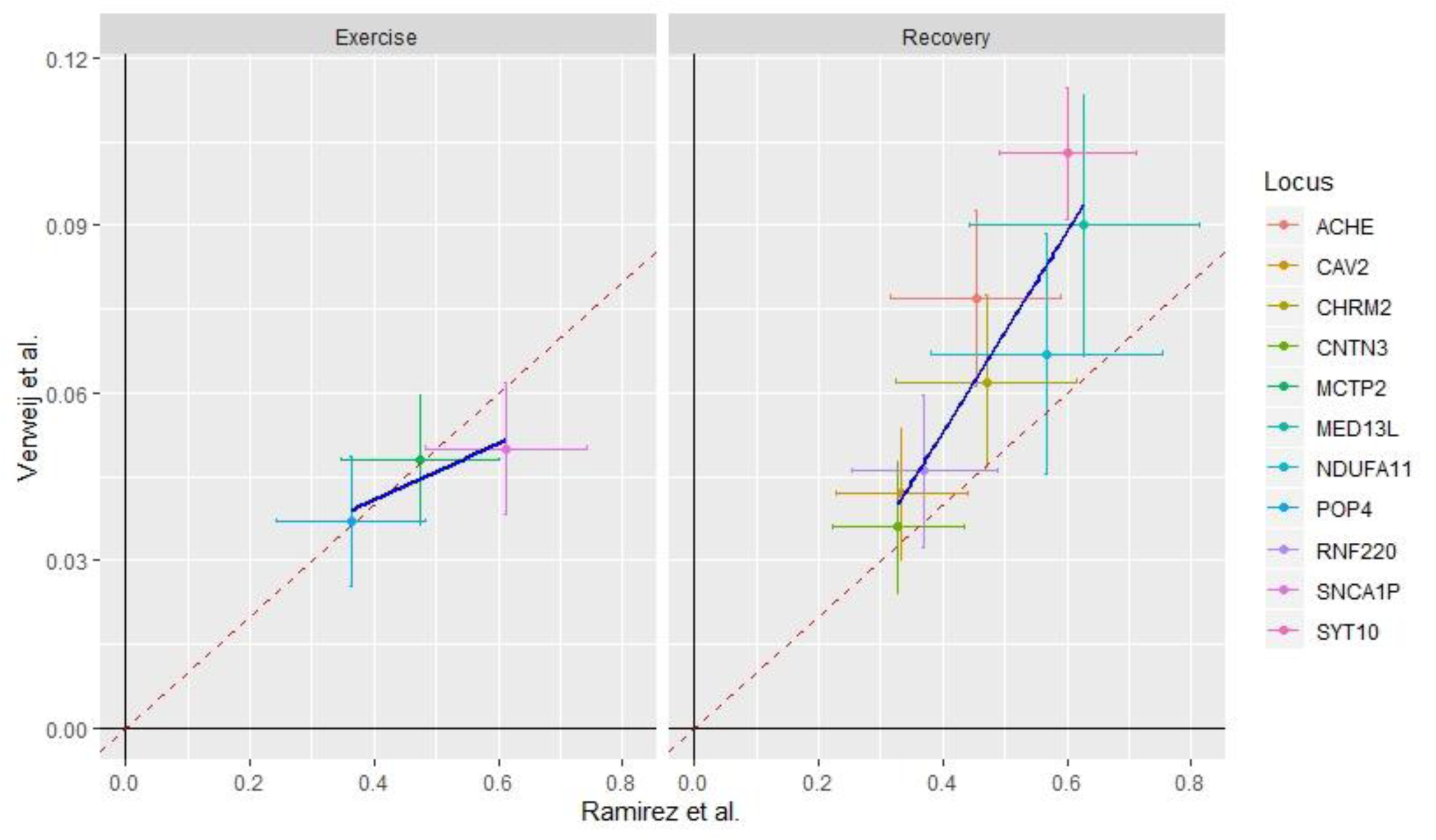
Comparison of the estimates and confidence intervals for the common SNPs for the two studies included I the review. The X-axis shows the estimates from the Ramirez et al. study and the Y-axis shows the estimates from the Verweij et al. study. The blue line is the regression line, and the dotted red line is the line of equivalence. The colors indicate the loci within the vicinity of the SNPs.

The beta estimates for Verweij et al. are for the inverse normal transformed traits. On an average, the beta estimates computed by Verweij et al. are 0.14 times those computed by Ramirez et al. Further, all the effects of the SNPs are in the same direction in both the analysis and the ranks are similar, taking strand effects into account (Figure 1).

Supplementary Table 1 provides a comparison of the results of the two studies. This table allows for the comparison of the SNP effects and examines the linkage disequilibrium between different SNPs described by the two studies at the same loci.

In terms of the PRS, Ramirez et al. developed scores in the full dataset using the beta estimates of the lead and secondary SNPs from the replication analysis, presumably after adjusting the estimates of the primary SNPs for the secondary SNPs after conditional analysis. They then standardized the score to have a mean of zero and a standard deviation of one. On the other hand, Verweij et al., generated multiple scores, one for each of the six traits analyzed, as well as a score that included SNPs across traits. These scores were based on the beta coefficients of all the primary and secondary SNPs, which were genome-wide significantly associated. Verweij et al. did not standardize their scores; instead, they tested the score in an independent dataset of the UK Biobank that did not participate in the cardio assessment. In this test, alleles were weighted with the effect on the highest associated HR-response phenotype (15); they found that this score was significantly associated with diastolic blood pressure(include beta, SE, P= 2×10^-25^) and pulse pressures (include beta, SE, P=3×10^-15^). Higher scores were associated with lower diastolic and pulse pressures. However, this score was not significantly associated with systolic blood pressure (include beta, SE, P=0.63).

A comparison of the distribution of the polygenic risk scores standardized (to means of zero and SD of 1) in the 1000 genomes European super-population is presented in Supplementary Figure 3. Scatterplots of the standardized scores are presented in Supplementary Figure 4. There was a good correlation between standardized study scores. The correlation coefficients of the scores of the different traits within and between studies are presented in Supplementary Table 2. Bland-Altman plots are presented in Supplementary Figure 5. A Bland-Altman plot is an analytic technique frequently used in clinical chemistry. It plots the mean of a pair of observations on the X-axis and their differences on the Y-axis. There is a good agreement of the observations if >95.4% of the values lie within ±2 SD of the mean(26). As is evident from the figure, there was a good agreement between the scores.

## 4. Discussion

Our systematic literature search revealed two large GWAS of short-term changes in HR during and after exercise. To our knowledge, this is the first review of the polygenic scores of short-term changes in HR during and after exercise, based on large sample sizes. Both studies in the review were based on the UK Biobank. There were several similarities in their methodologies. Indeed, the two studies discovered 13 SNPs that were common for the recovery traits and 7 SNPs that were common to exercise traits. However, there were several differences in their findings, which were presumably due to the different methodologies followed by the two groups.

There are several significant differences in the methodologies used by the two groups. Firstly, the analyzed populations in the two studies had essential differences. These differences were due to different exclusion criteria, focus on different ancestries, and different algorithms to ascertain the discrepancies between the UK Biobank provided heart rates and ECG recordings. Secondly, while Ramirez et al. split their dataset into discovery (n∼40k) and replication subsets (n∼27k), Verweij et al. analyzed the whole dataset together (n∼58k). Thirdly, the two studies defined recovery-related traits differently. While Ramirez et al. conducted one GWAS for their recovery trait, Verweij et al. conducted five GWAS for their five recovery traits. Fourthly, the two studies used different covariates in their analyses. Finally, Verweij et al. used inverse-normal transformation before the analysis, while Ramirez et al. analyzed raw traits.

The analytic approach followed by Verweij et al. could have led them to miss some associations. The approach assumed independence of the five recovery traits and, therefore, employed stringent significance criteria for association with the SNPs. However, the same authors found that the traits are phenotypically and genetically correlated (15). Further, there was a strong correlation of the Verweij recovery PRS (see Supplementary Table 2) in the 1000 Genomes European super-population. Thus, the stringent criteria used for testing the significance of association may have been unjustified, leading to an erroneous rejection of some associations (Type 2 error).

We suggest two possible alternatives for analyzing correlated heart rate recovery traits. Firstly, instead of performing five separate GWAS for recovery traits, one could employ the linear mixed models (LMM) approach to take advantage of repeated observations at different points in time. Although linear mixed models for GWAS are computationally challenging, a recent algorithm circumvents many of these challenges (31). Secondly, one could use a multivariate approach. This approach simultaneously tests the association of linear combinations of multiple phenotypes with genotypes at each SNP and could provide a significant increase in power (32).

In both the studies, there is a potential for selection bias operating at different levels. Firstly, there is potential for bias at the level of participation in the UK Biobank. The participants in the UK Biobank are more likely to be healthier and wealthier than non-participants (33). Secondly, of the UK Biobank participants, only 100,000 were selected for exercise testing(27). The selection appears to be random, although a comparison between those selected and those not selected would be required to justify this statement. Thirdly, of those selected for participation in exercise testing, only a few more than 90,000 completed the full protocol (34). The rest could not complete the protocol because of various reasons, including wanting to stop early, developing chest pain or discomfort, heart rate reaching safety level, amongst others (34). The fourth level of potential selection bias operated only for Verweij et al. They excluded people with high cardiovascular risk. Finally, the exclusion of non-Europeans by Ramirez et al. could potentially be a source of another bias.

Another source of possible difference between the studies could be the rank-based inverse normal transformation of traits performed by Verweij et al. They justified this transformation by stating that it improves the power to detect associations with low-frequency variants and improves the comparability between traits (15). However, simulations of this transformation have shown that it may neither maintain type-1 error control nor have a power advantage over other methods (35). Despite the differences in the transformation of outcomes, the ordering of the associations between the two studies are comparable (Figure 1).

Our review presents several insights that are important to human health. The review identifies the SNPs that are associated with short-term changes in HR during and after exercise, traits which can be used to assess the cardiovascular health of individuals. The knowledge of the identified SNPs will help in fueling studies aimed at identifying the cellular and molecular pathways that underlie these responses. This knowledge will also help in identifying targets for pharmacotherapy for cardiovascular disorders. Further, the PRS could be used in the analysis of correlated traits and to predict cardiovascular health and mortality from cardiovascular diseases. The utility of PRS for predicting mortality from cardiovascular diseases has been assessed by Ramirez et al. (14). However, their analysis was not sufficiently powered owing to the number of events in the observed timeframe.

Both studies used polygenic scores derived from genome-wide significant SNPs. However, previous research has shown that the polygenic scores derived from all SNPs perform better in terms of prediction of outcomes as opposed to those derived by including only genome-wide significant SNPs (17).

Although a previous report (36) has reviewed the two studies, they summarized the findings from the two studies as a Venn diagram without a mention of the magnitude and direction of the effect. Further, the Venn diagram of the review was difficult to reproduce. A second review (37) also summarized these two studies. However, that review combined the results of the two studies with several candidate gene studies and GWAS with smaller sample sizes. Thus, bias due to the winner’s curse is likely in this review. Further, both of these reports (36, 37) have not commented on the polygenic scores derived from the studies.

This review has several strengths. Firstly, it compares two studies on subsets of the UK Biobank including analyzes how different approaches by different groups answer different on similar datasets. Secondly, the present review systematically assesses the completeness of reporting of evidence for the associations of SNPs with the HR traits based on robust guidance(22), especially in the context of biases common to GWAS (winner’s curse). Thus, it emphasizes the importance of replication.

Despite the strengths of the review, there are several limitations. Firstly, despite a systematic approach, we could only identify two GWAS of short-term changes in HR during and after exercise. Because of the overlap in the subsets analyzed by the identified studies, we could not perform a meta-analysis. Improving the effect size estimates is an essential goal of replication. Secondly, the included studies were both performed on the UK Biobank. This Biobank has representation from the UK population. Although there is a representation of individuals of different ancestries, the Biobank includes individuals of predominantly European ancestry (16). Further, Ramirez et al. only analyzed data on individuals of European ancestry. Thus, the review lacks a representation of data on individuals of different ancestries.

Future studies would need to address the gap of lack of data on individuals of under-represented ancestries.

## 5. Conclusion

GWAS are one of the most powerful tools to assess the strength of association between genetic polymorphisms and traits. GWAS can be used to develop PRS. The performance of these PRS improves with large sample sizes of the GWAS from which they are derived. Our systematic literature review found two GWAS of short-term changes in HR with exercise with more than 10,000 participants. Both studies were conducted on the UK Biobank and found several common SNPs and several others that were mutually exclusive. There was a good agreement in the polygenic scores from the two studies. The difference in methodologies adopted by the two groups and their findings highlight the importance of replication in the scientific process. The underrepresentation of individuals of non-European ancestry in genetic studies is a challenge to the generalization of their findings. Future studies with a proportionate representation of individuals of non-European ancestry would be required to address this gap.

## Data Availability

The datasets were derived from sources in the public domain and are available from the following URLs:
LDLINK: https://ldlink.nci.nih.gov; LDlink 4.1.0 Release (04/29/2020), accessed date: 2020 May 20th
1000 Genomes: https://www.internationalgenome.org/data; Accessed 2020, June 3rd
UK Biobank: http://biobank.ndph.ox.ac.uk/showcase/field.cgi?id=6020 ; Accessed 2020 August 6
th

## 7. Data Availability Statement

The datasets were derived from sources in the public domain and are available from the following URLs: LDLINK: https://ldlink.nci.nih.gov; **LDlink 4.1.0 Release (04/29/2020), accessed date: 2020 May 20^th^** 1000 Genomes: https://www.internationalgenome.org/data; **Accessed 2020, June 3^rd^** UK Biobank: http://biobank.ndph.ox.ac.uk/showcase/field.cgi?id=6020 ; **Accessed 2020 August 6^th^**

---

**Supplementary Figure 1.**
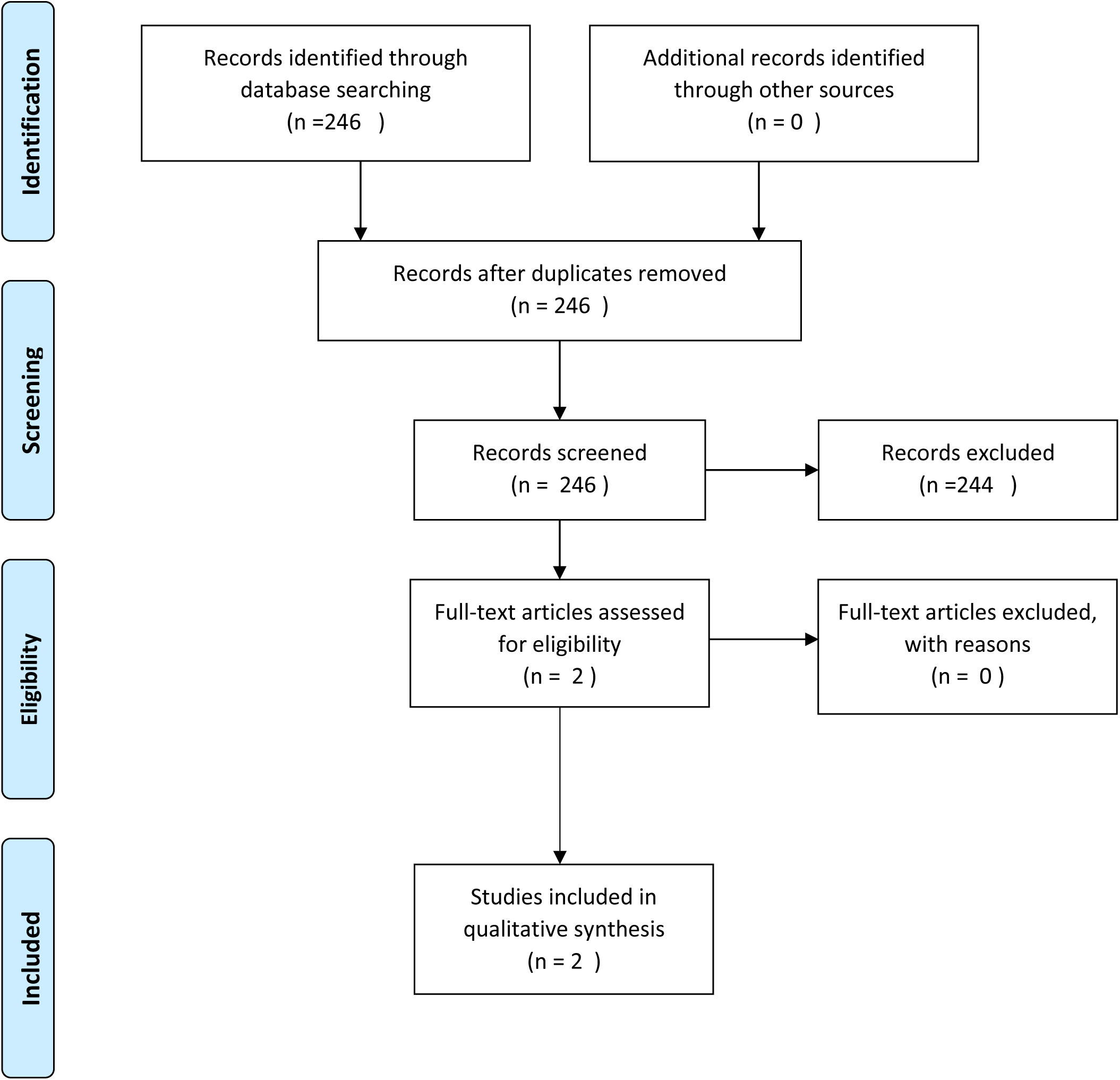
PRISMA Flow Diagram for the Search Strategy

**Supplementary Table 1:**
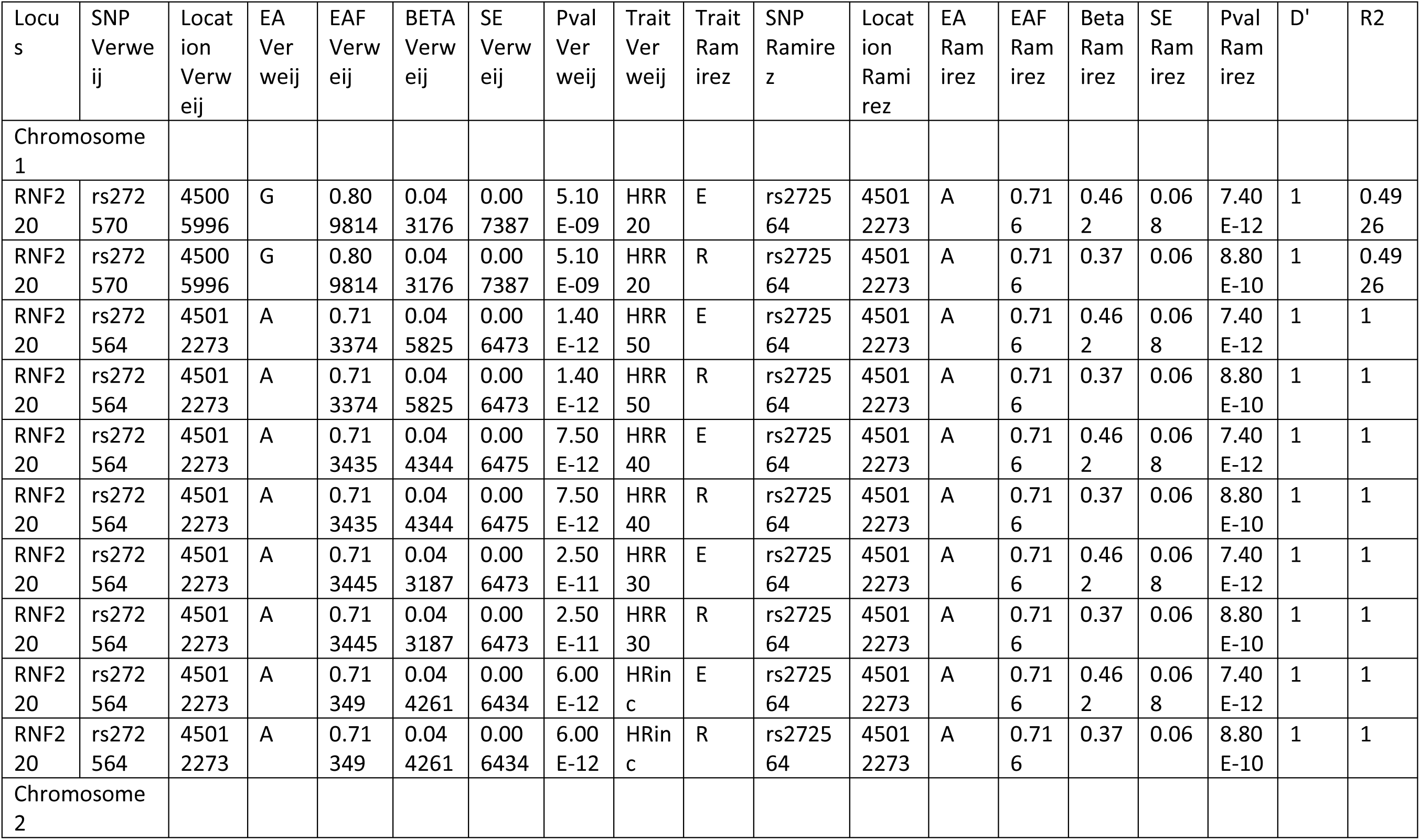
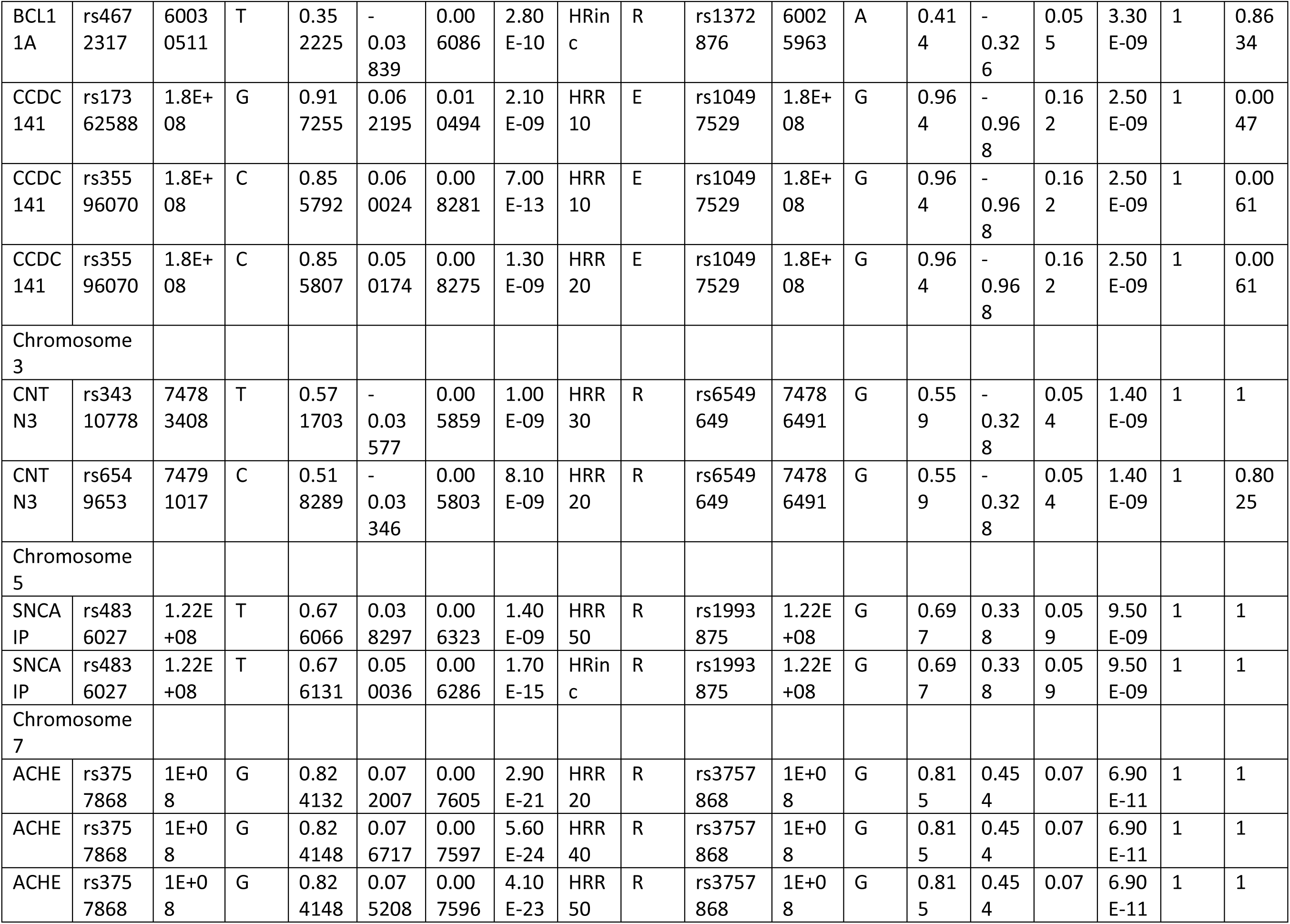
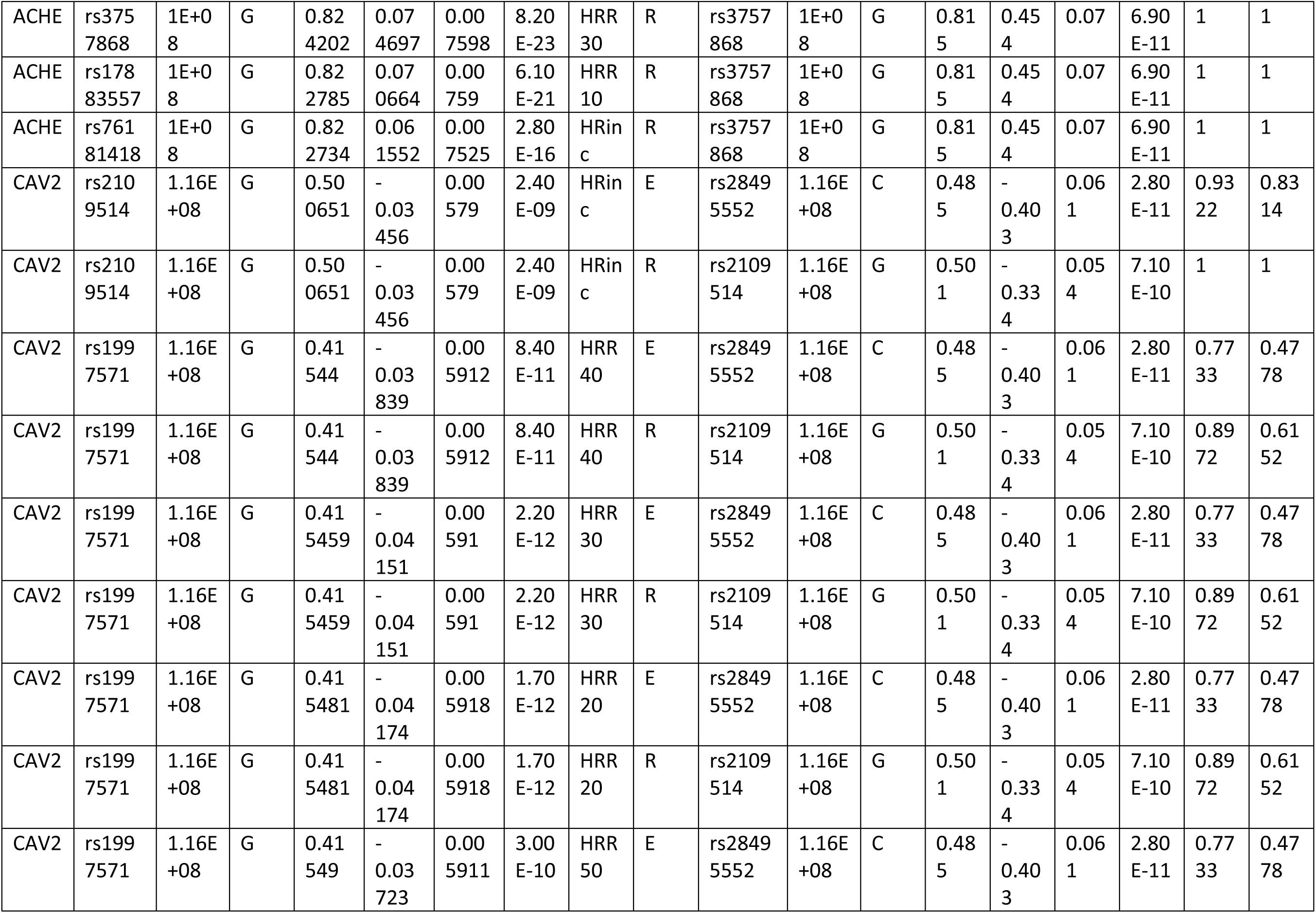
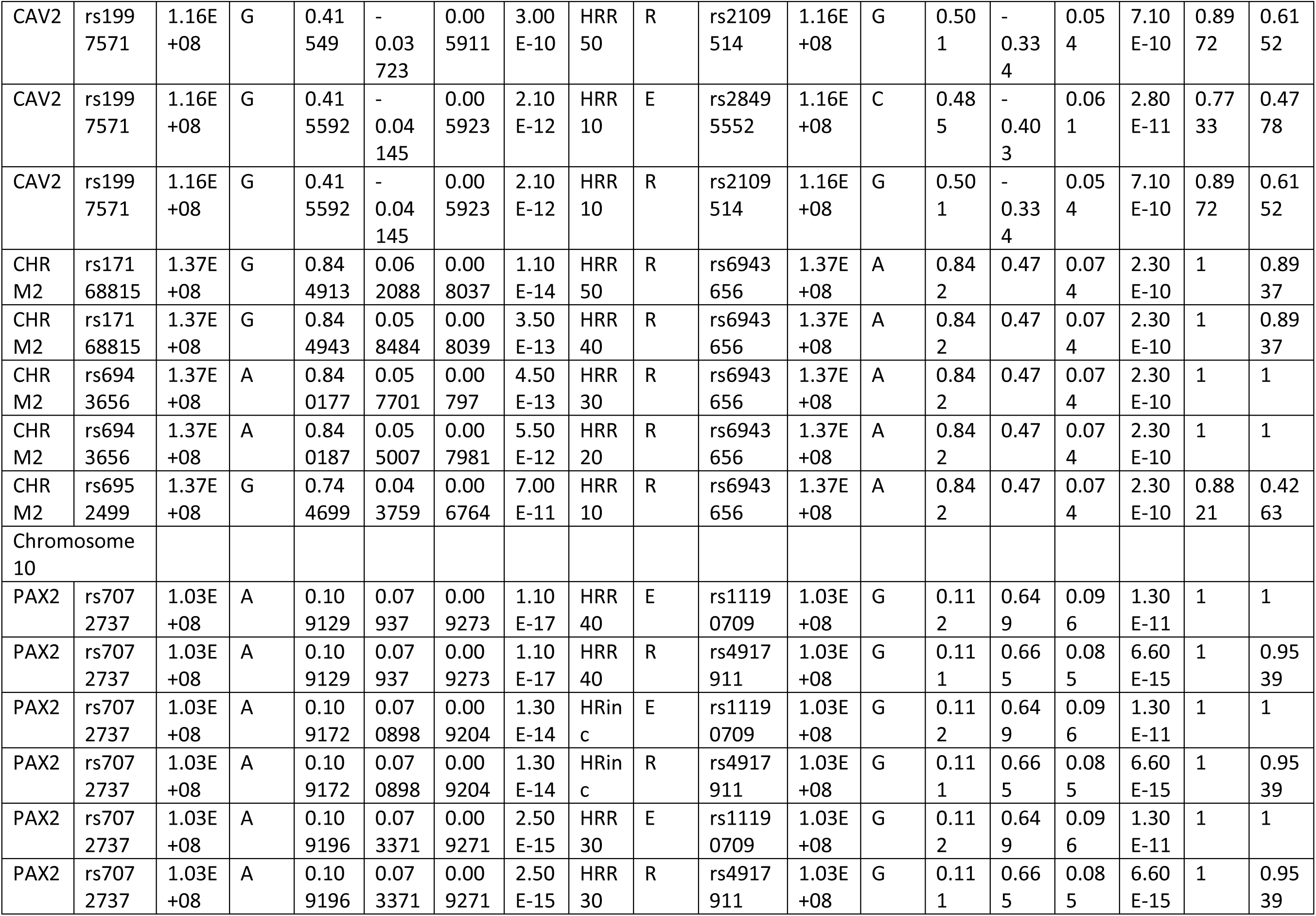
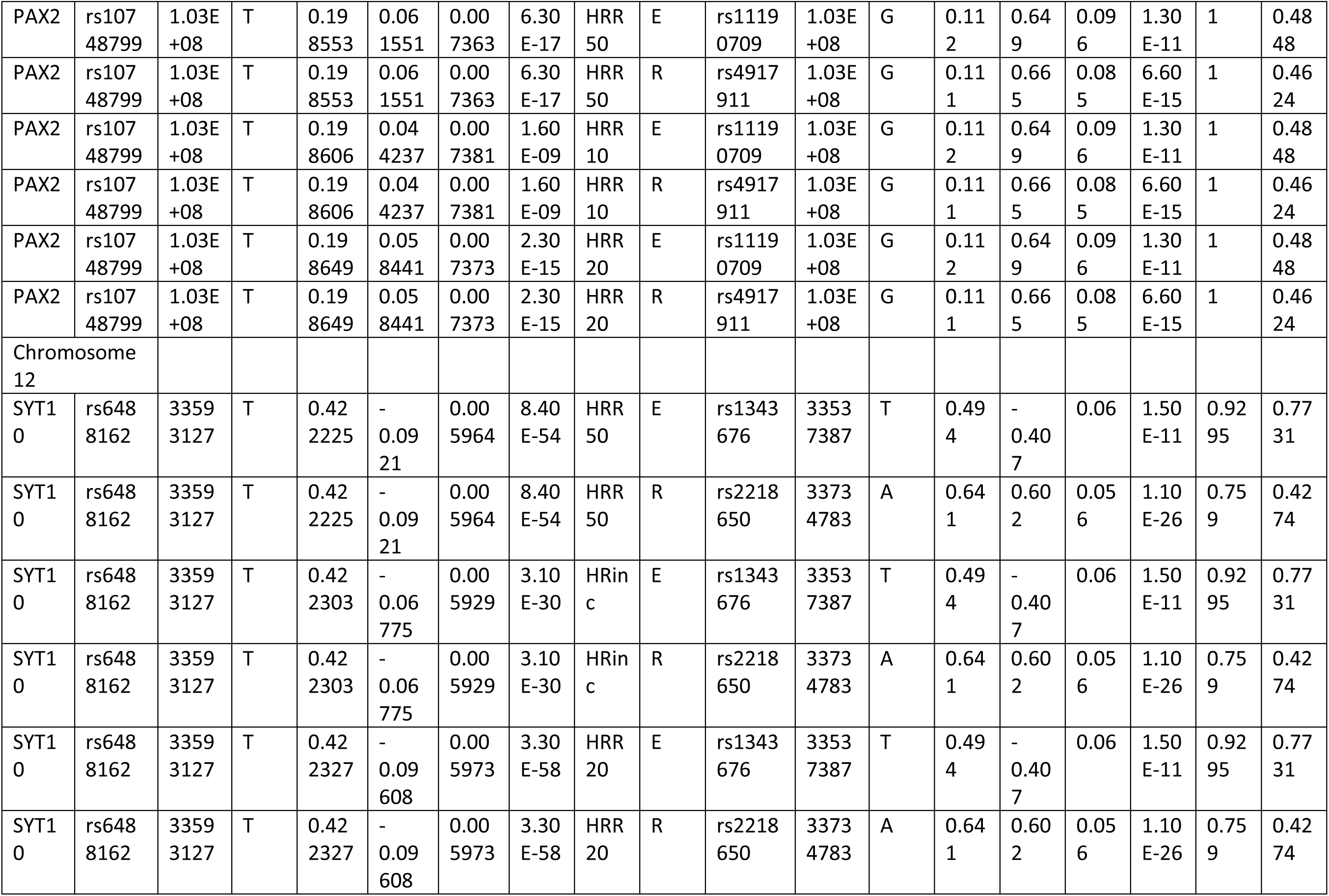
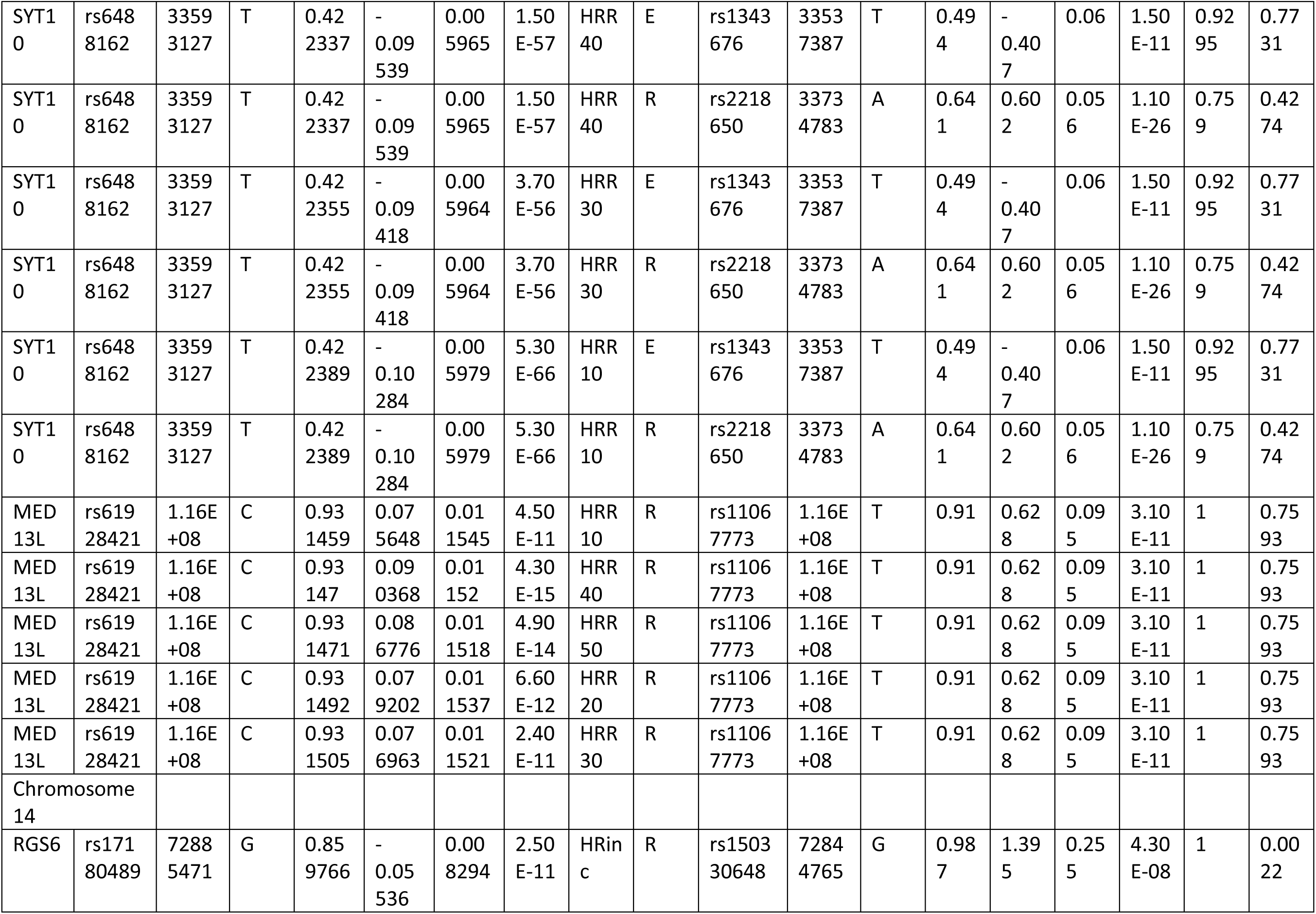
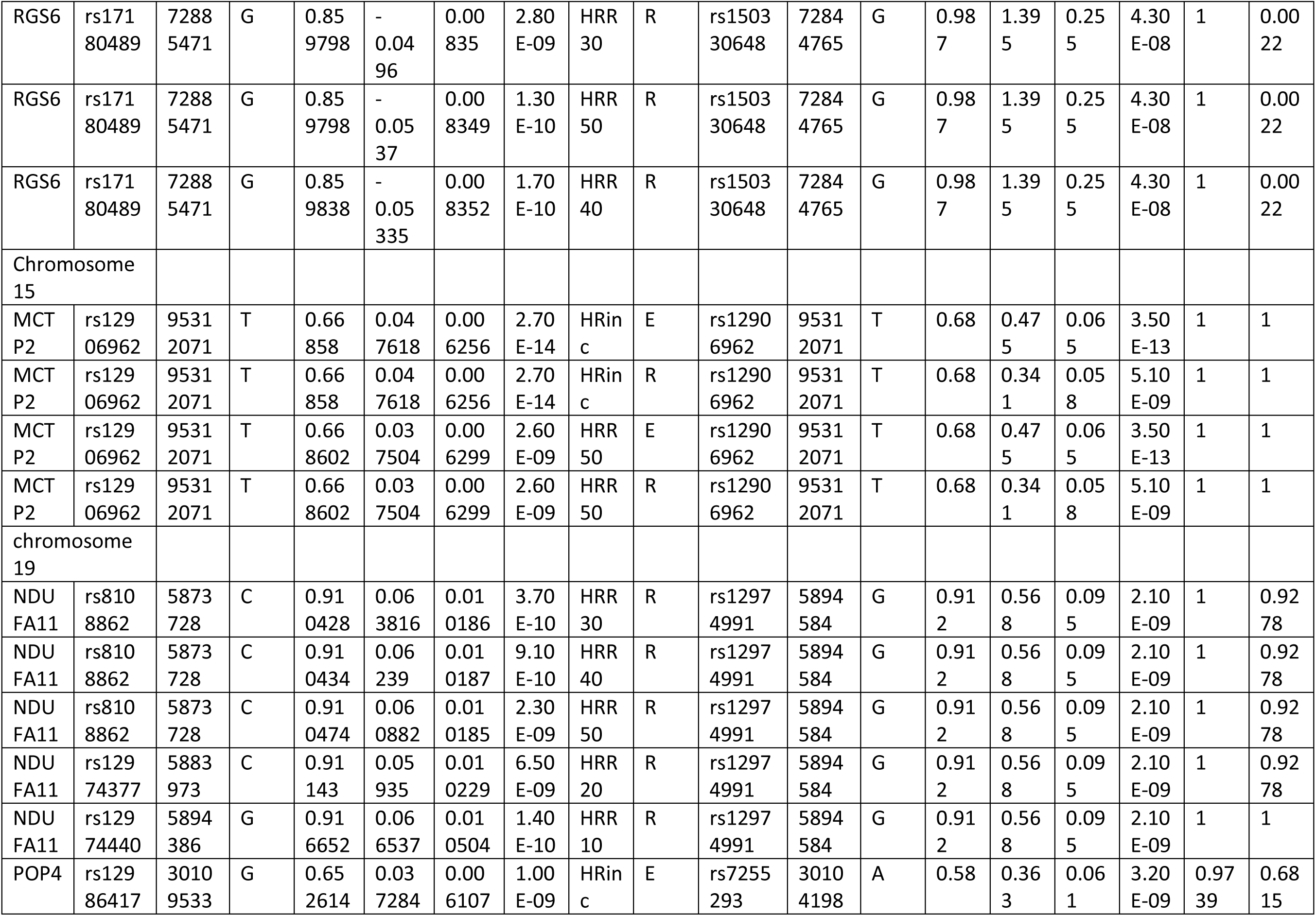
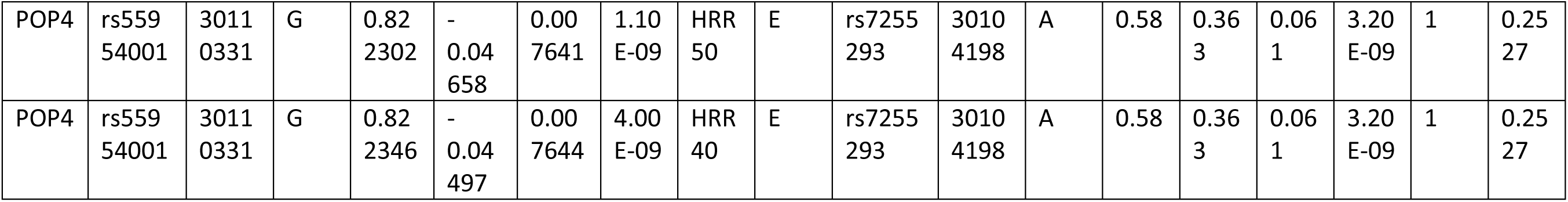
Comparison of Loci Common to the Two Studies. EA = Effect Allele. EAF= effect Allele Frequency. BETA= Beta estimate, SE= Standard Error. Pval= P value. D’ and R2 are from the GBR population of the 1000 Genomes population. E= Heart Rate increase with Exercise in Ramirez & Verweij. R= Heart rate Recovery Trait In Ramirez. HRR= Heart Rate Recovery Trait for 10-50 second in Verweij study

**Supplementary Table 2:** Correlation between the Polygenic scores in the European superpopulation of 1000 genome for traits between and within studies. Values above the diagonal are correlation coefficients. Values below the diagonal are P values.

| Traits | Ramirez Exercise | Ramirez Recovery | Verweij Exercise | Verweij Recovery 10s | Verweij Recovery 20s | Verweij Recovery 30s | Verweij Recovery 40s | Verweij Recovery 50s |
| --- | --- | --- | --- | --- | --- | --- | --- | --- |
| Ramirez Exercise |  | 0.54 | 0.59 | 0.12 | 0.11 | 0.16 | 0.26 | 0.40 |
| Ramirez Recovery | 1.90E-39 |  | 0.45 | 0.08 | 0.10 | 0.25 | 0.40 | 0.40 |
| Verweij Exercise | 1.07E-48 | 7.65E-27 |  | 0.03 | 0.03 | 0.13 | 0.2 | 0.32 |
| Verweij Recovery 10s | 0.007 | 0.06 | 0.460 |  | 0.80 | 0.61 | 0.61 | 0.61 |
| Verweij Recovery 20s | 0.010 | 0.022 | 0.494 | 8.25E-114 |  | 0.76 | 0.67 | 0.65 |
| Verweij Recovery 30s | 2.67E-04 | 1.54E-08 | 0.002 | 1.14E-51 | 5.08E-97 |  | 0.85 | 0.70 |
| Verweij Recovery 40s | 1.07E-09 | 1.85E-20 | 7.71E-06 | 5.33E-53 | 2.99E-66 | 4.54E-139 |  | 0.81 |
| Verweij Recovery 50s | 8.69E-21 | 8.06E-21 | 2.40E-13 | 1.73E-52 | 7.11E-62 | 2.72E-76 | 5.32E-119 |  |

**Supplementary Table 3:**
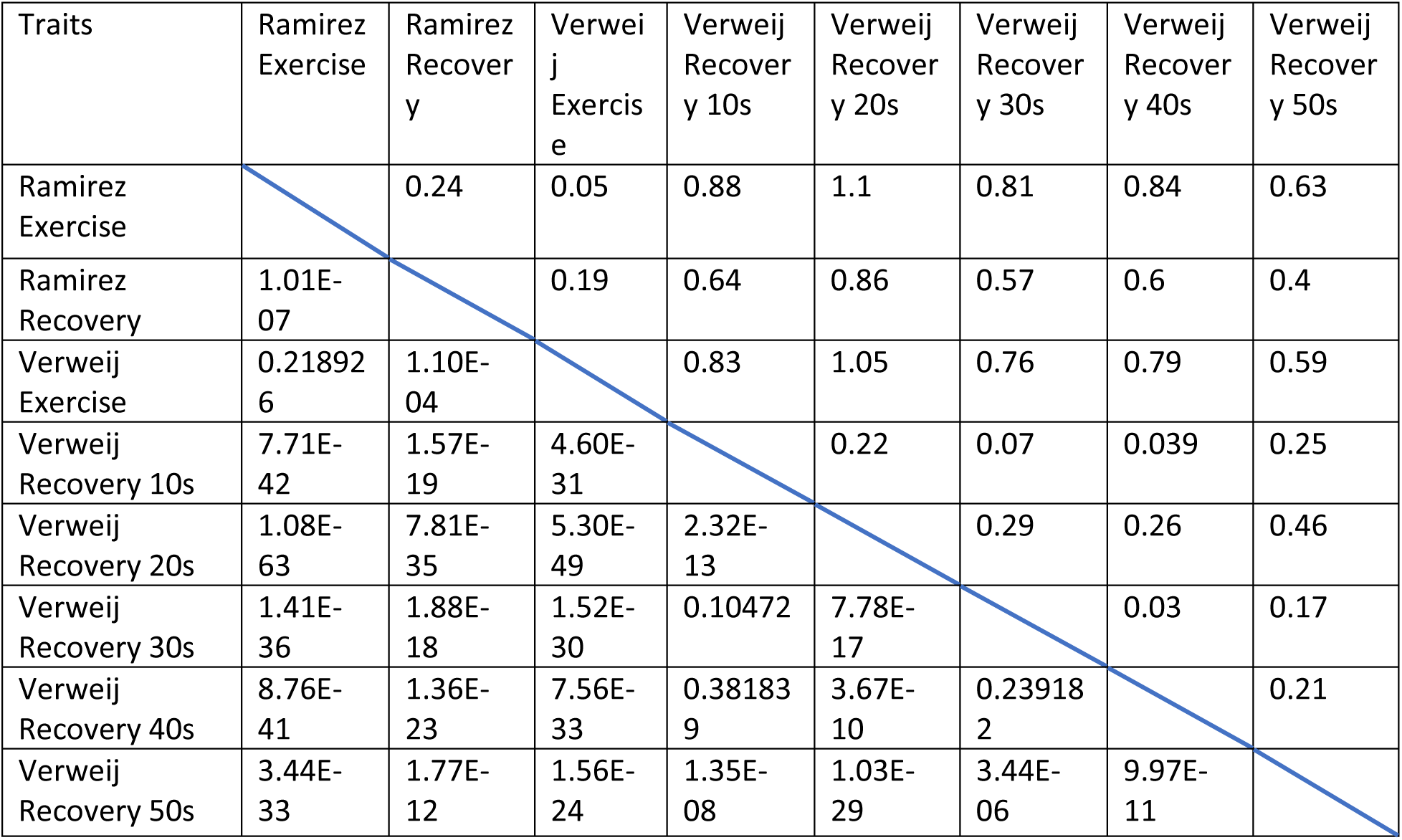
Mean Differences of the polygenic risk scores of exercise and recovery traits between and within studies in the European Superpopulation of 1000 Genomes. Values above the diagonal are means of differences. Values below the diagonal are P values.

| Traits | Ramirez Exercise | Ramirez Recovery | Verweij Exercise | Verweij Recovery 10s | Verweij Recovery 20s | Verweij Recovery 30s | Verweij Recovery 40s | Verweij Recovery 50s |
| --- | --- | --- | --- | --- | --- | --- | --- | --- |
| Ramirez Exercise |  | 0.24 | 0.05 | 0.88 | 1.1 | 0.81 | 0.84 | 0.63 |
| Ramirez Recovery | 1.01E-07 |  | 0.19 | 0.64 | 0.86 | 0.57 | 0.6 | 0.4 |
| Verweij Exercise | 0.218926 | 1.10E-04 |  | 0.83 | 1.05 | 0.76 | 0.79 | 0.59 |
| Verweij Recovery 10s | 7.71E-42 | 1.57E-19 | 4.60E-31 |  | 0.22 | 0.07 | 0.039 | 0.25 |
| Verweij Recovery 20s | 1.08E-63 | 7.81E-35 | 5.30E-49 | 2.32E-13 |  | 0.29 | 0.26 | 0.46 |
| Verweij Recovery 30s | 1.41E-36 | 1.88E-18 | 1.52E-30 | 0.10472 | 7.78E-17 |  | 0.03 | 0.17 |
| Verweij Recovery 40s | 8.76E-41 | 1.36E-23 | 7.56E-33 | 0.381839 | 3.67E-10 | 0.239182 |  | 0.21 |
| Verweij Recovery 50s | 3.44E-33 | 1.77E-12 | 1.56E-24 | 1.35E-08 | 1.03E-29 | 3.44E-06 | 9.97E-11 |  |

**Supplementary Figure 2A:**
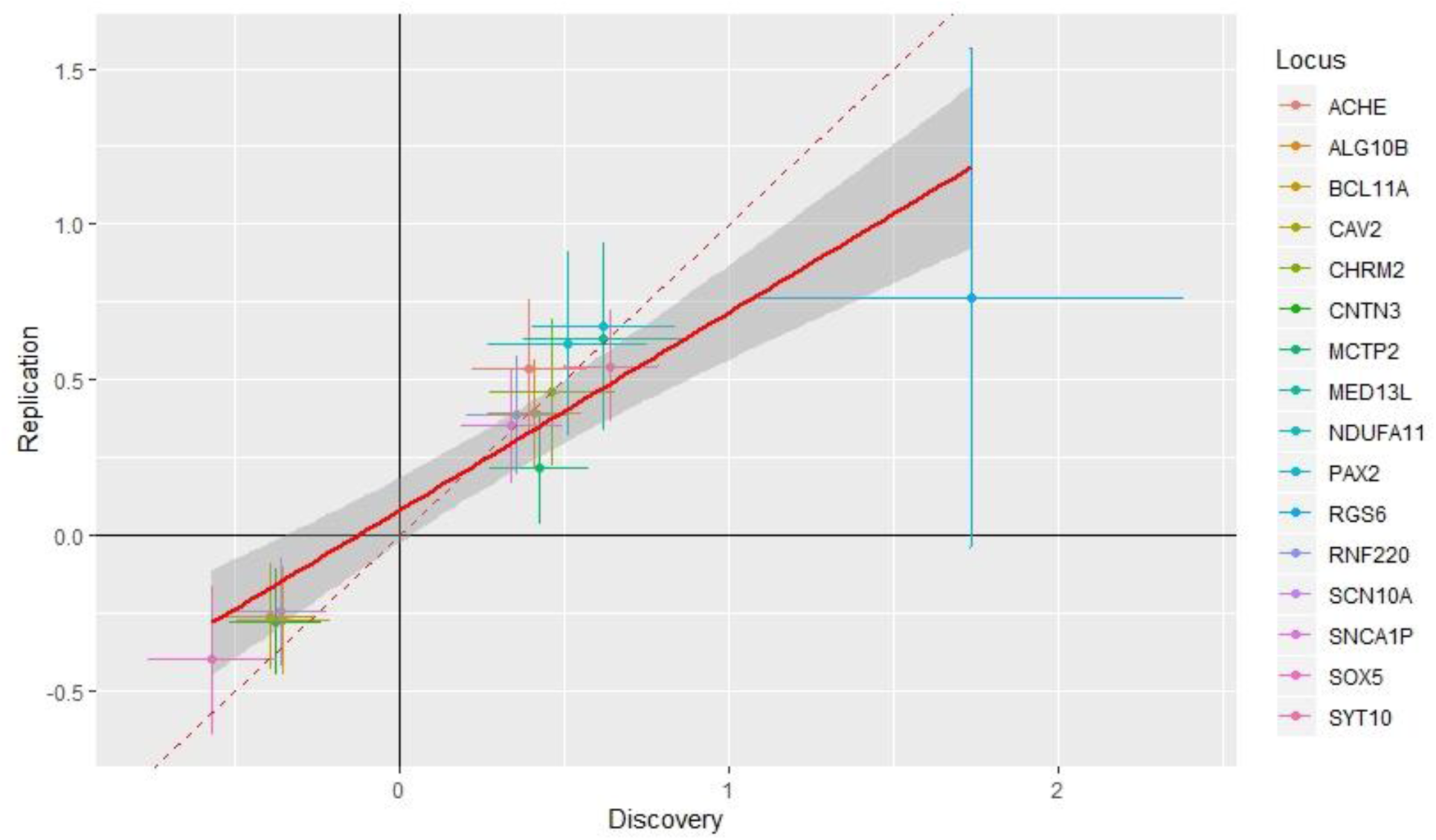
Comparison of beta estimates of Heart rate response to Recovery in the discovery and replication samples from Ramirez study

**Supplementary Figure 2B:**
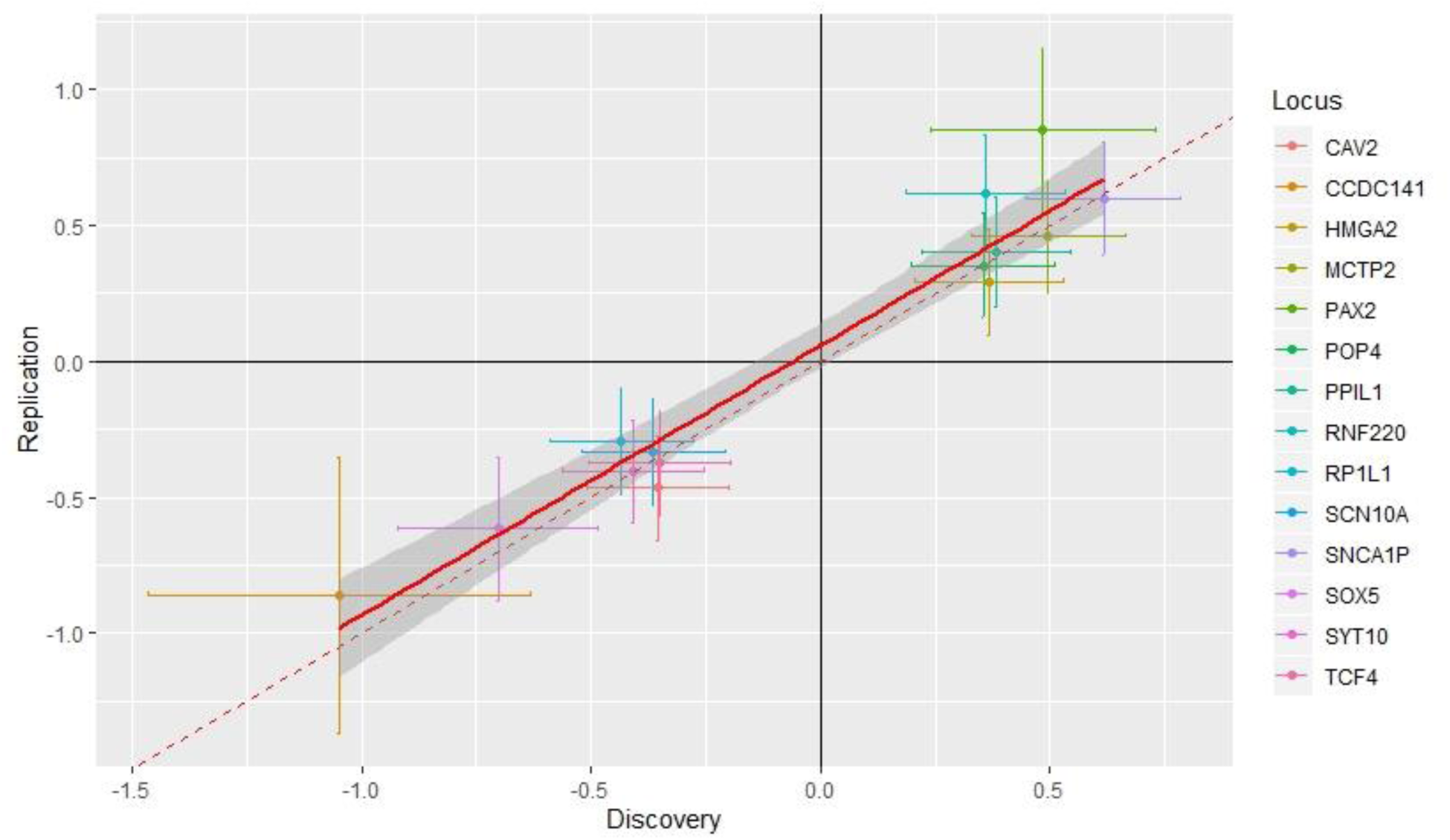
Comparison of beta estimates of Heart rate response to exercise in the discovery and replication samples from Ramirez study

**Supplementary Figure 2C:**
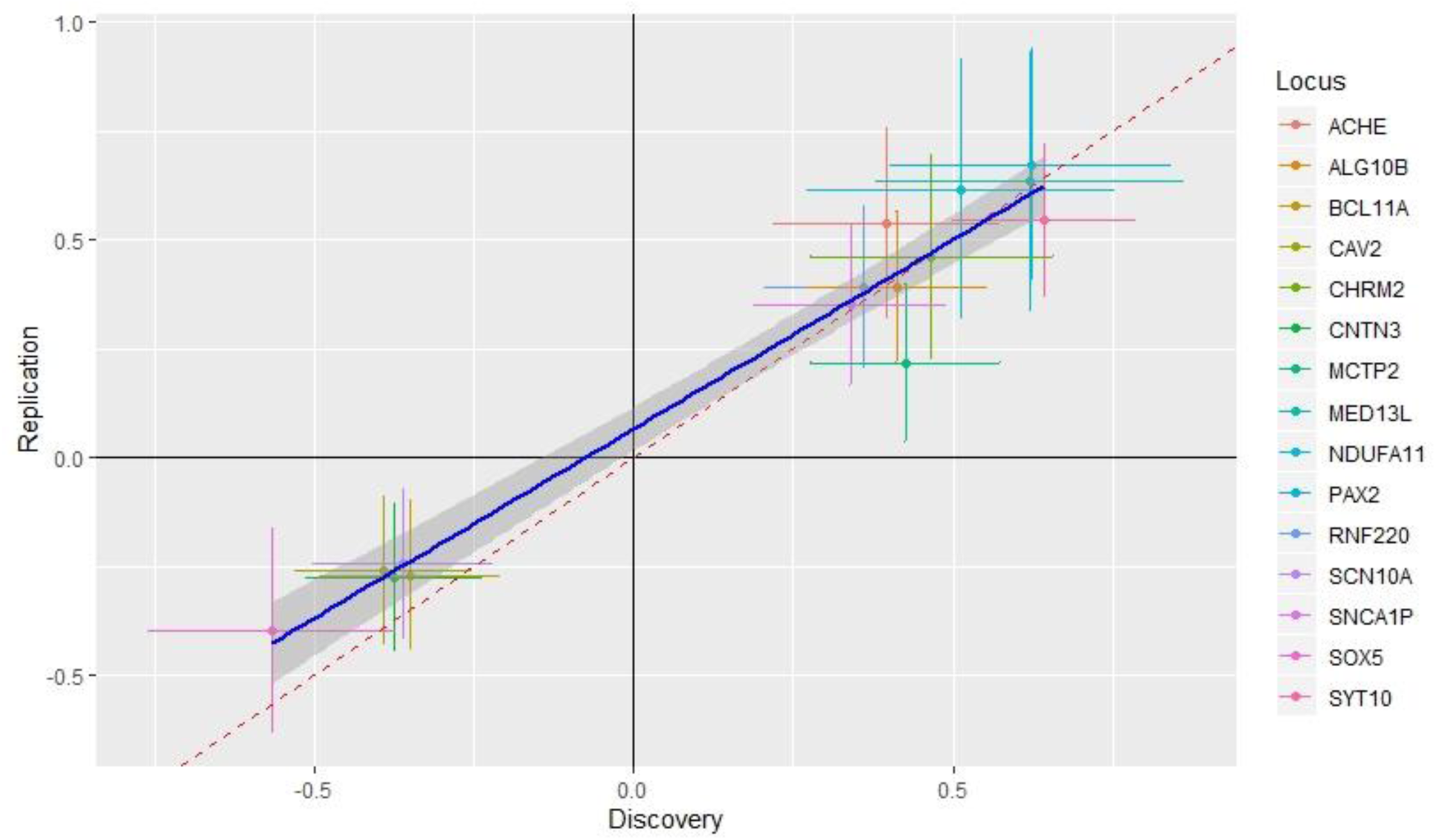
Comparison of beta estimates of Heart rate response to Recovery in the discovery and replication samples from Ramirez study after removal of the Outlier SNP

**Supplementary Figure 3A-3H:**
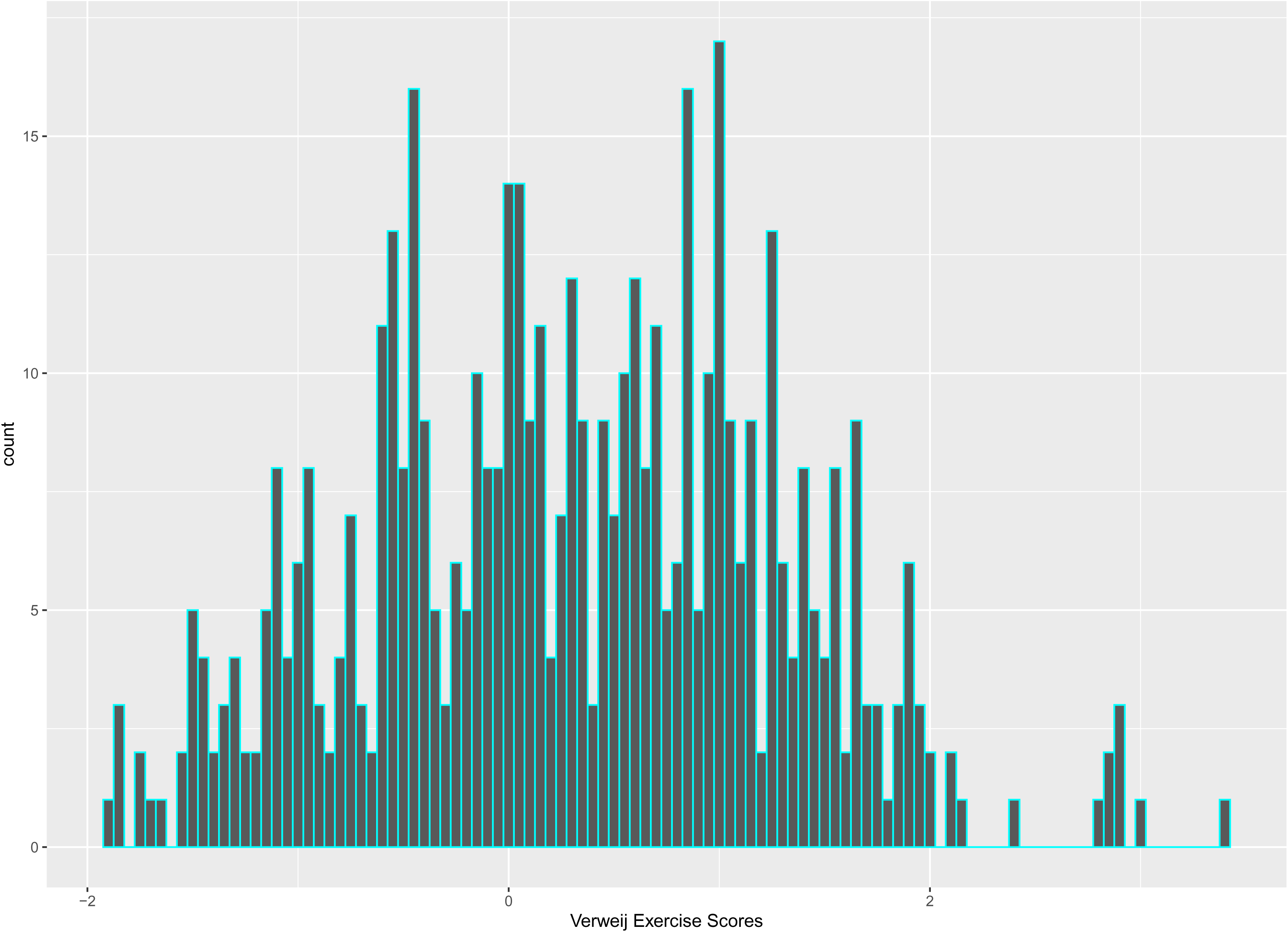
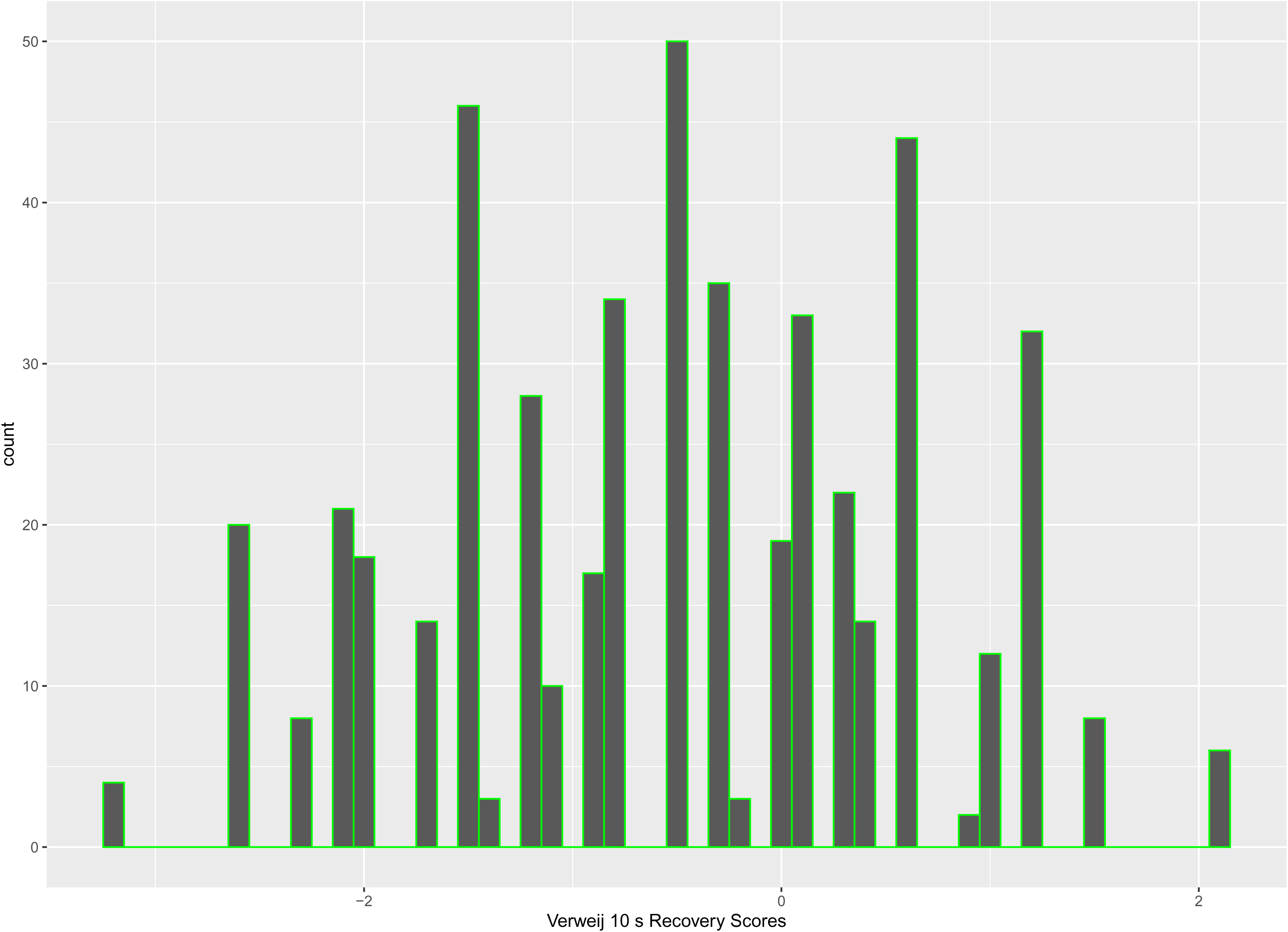
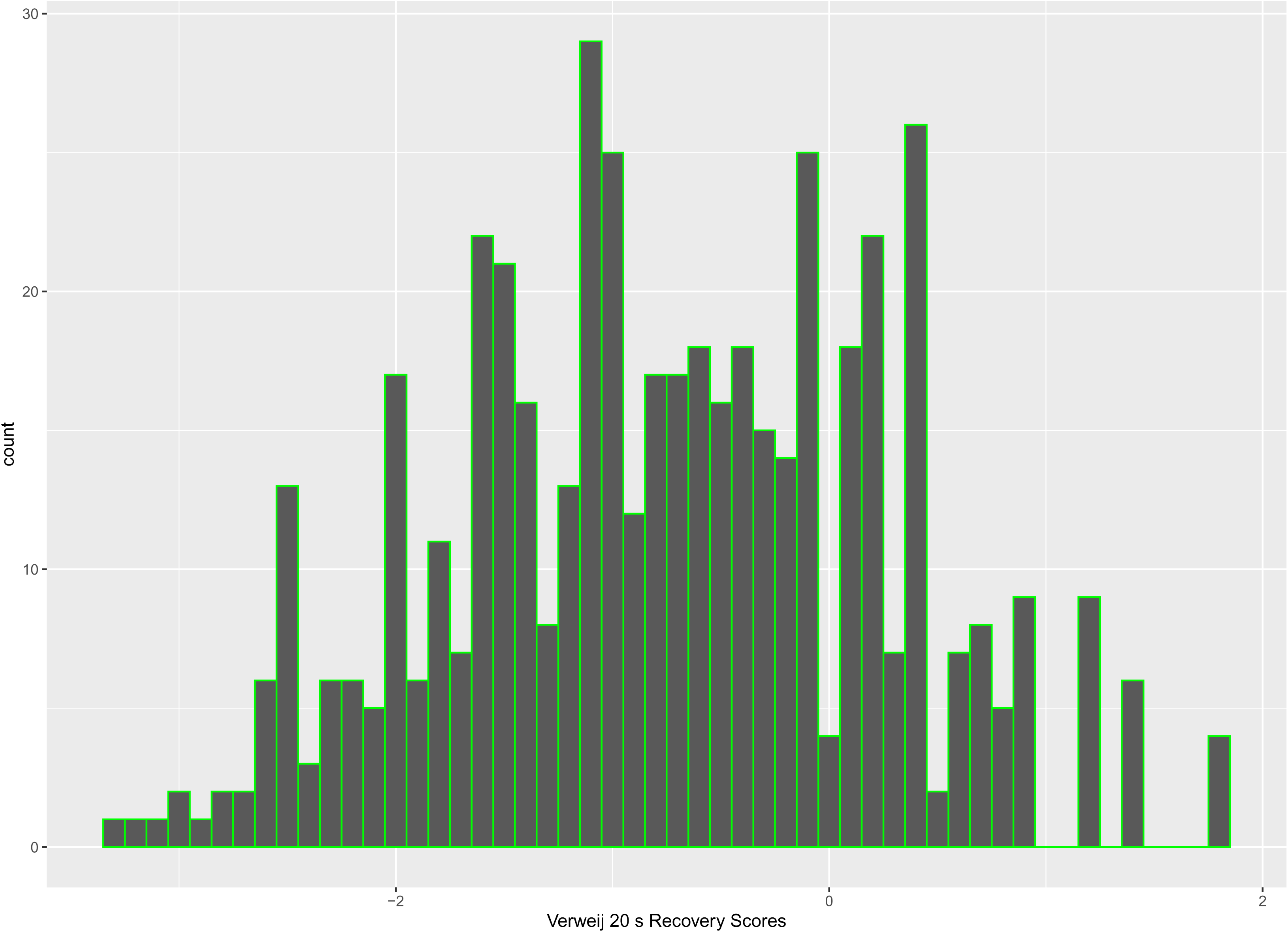
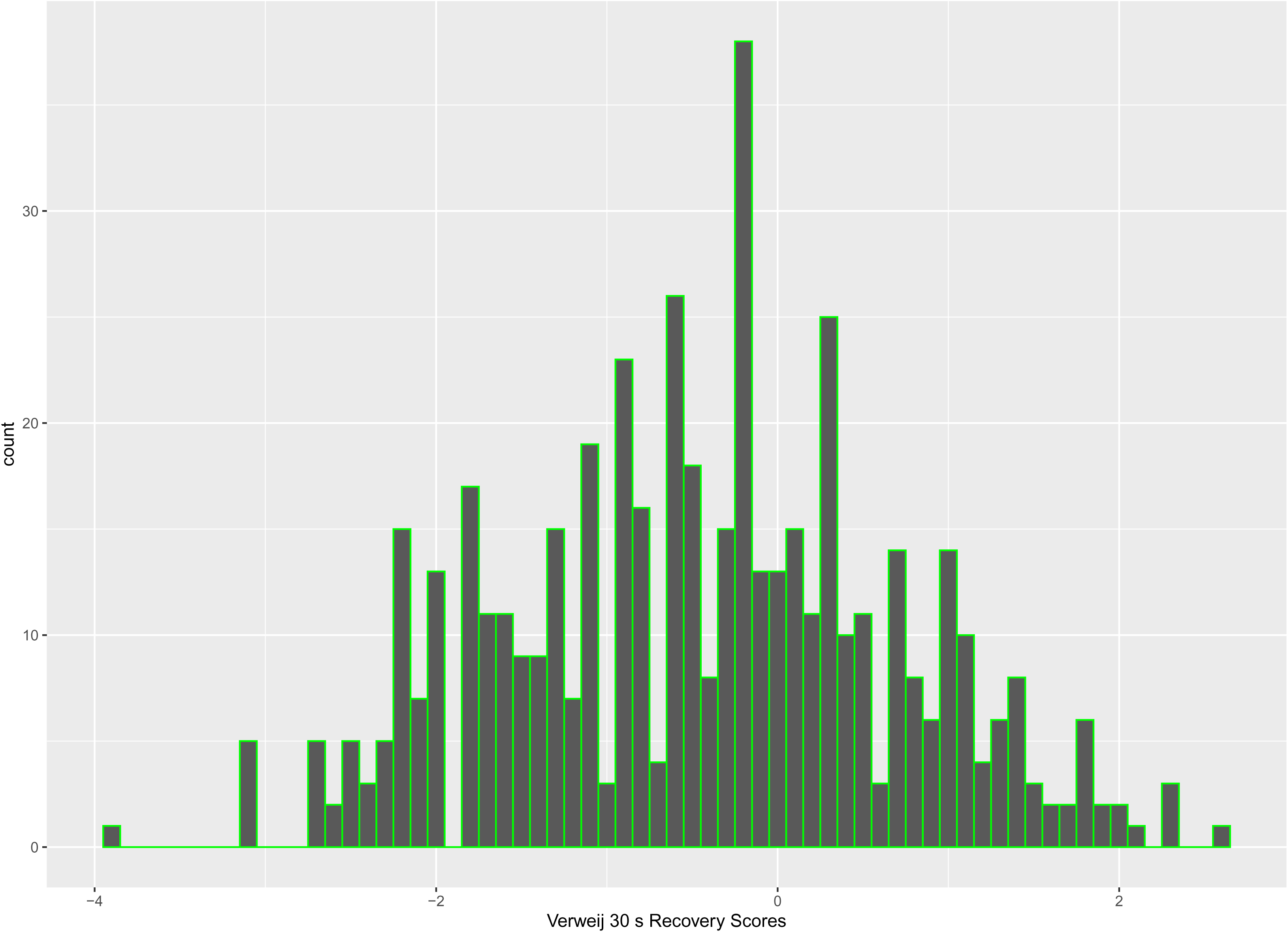
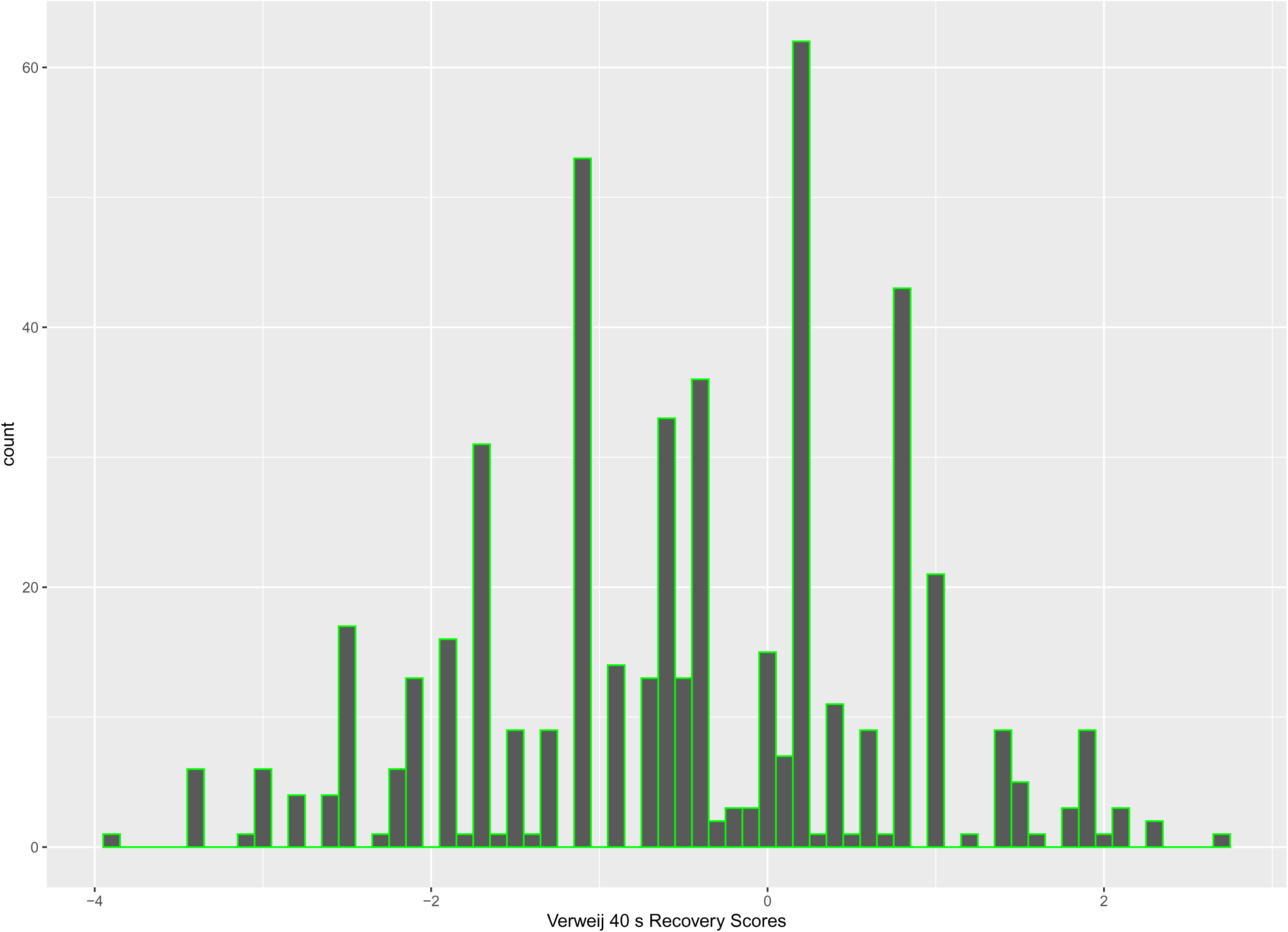
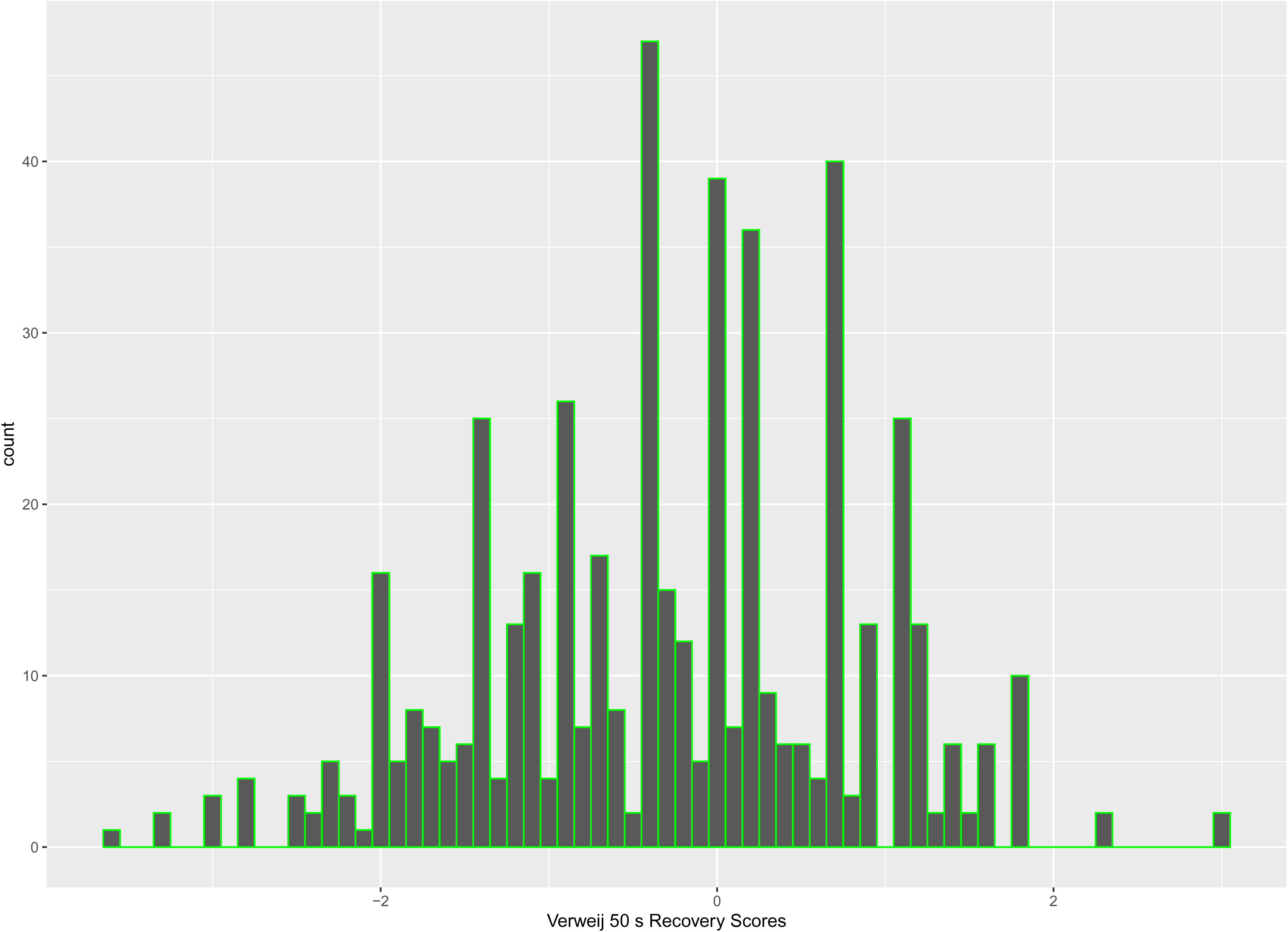
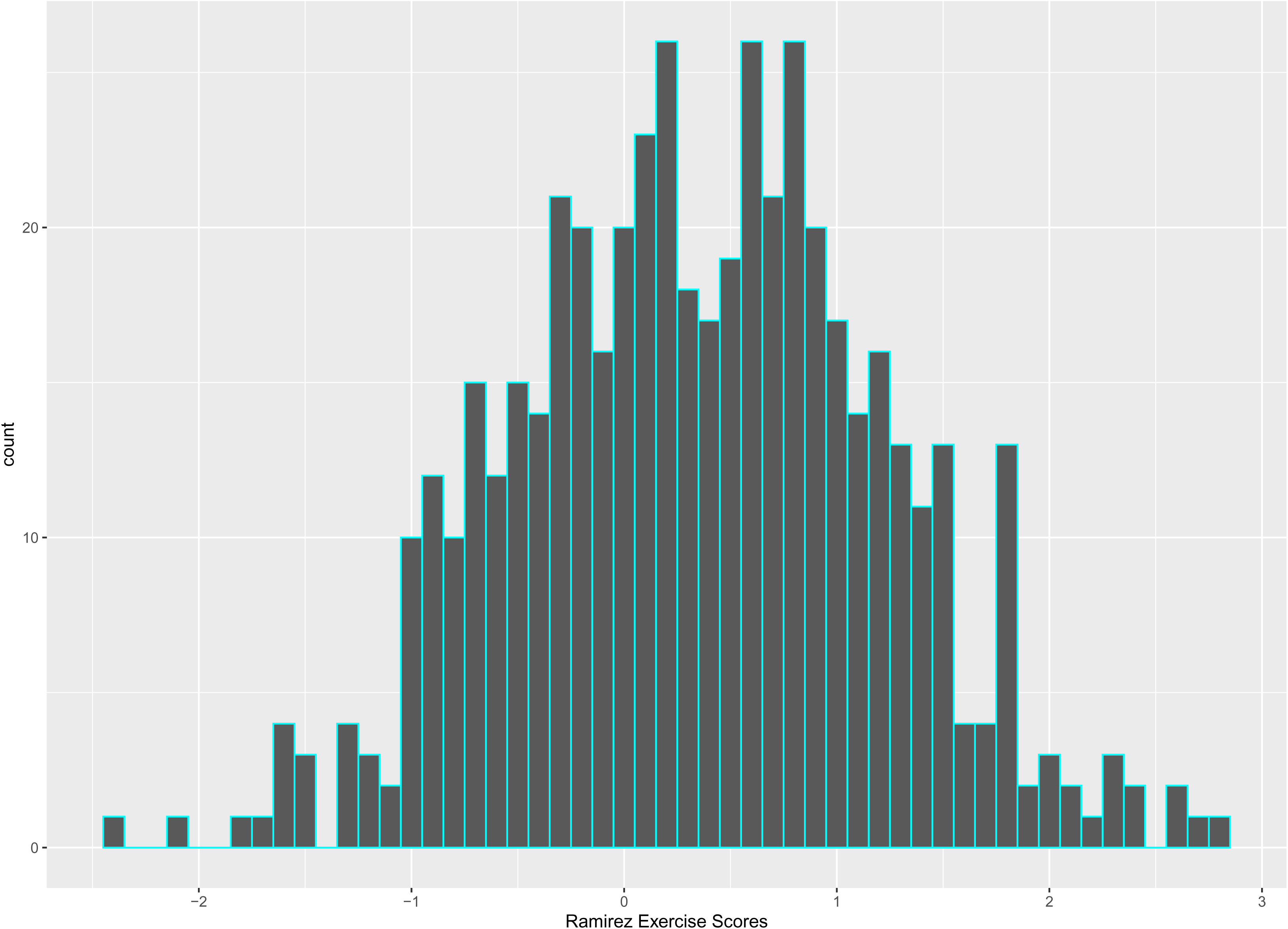
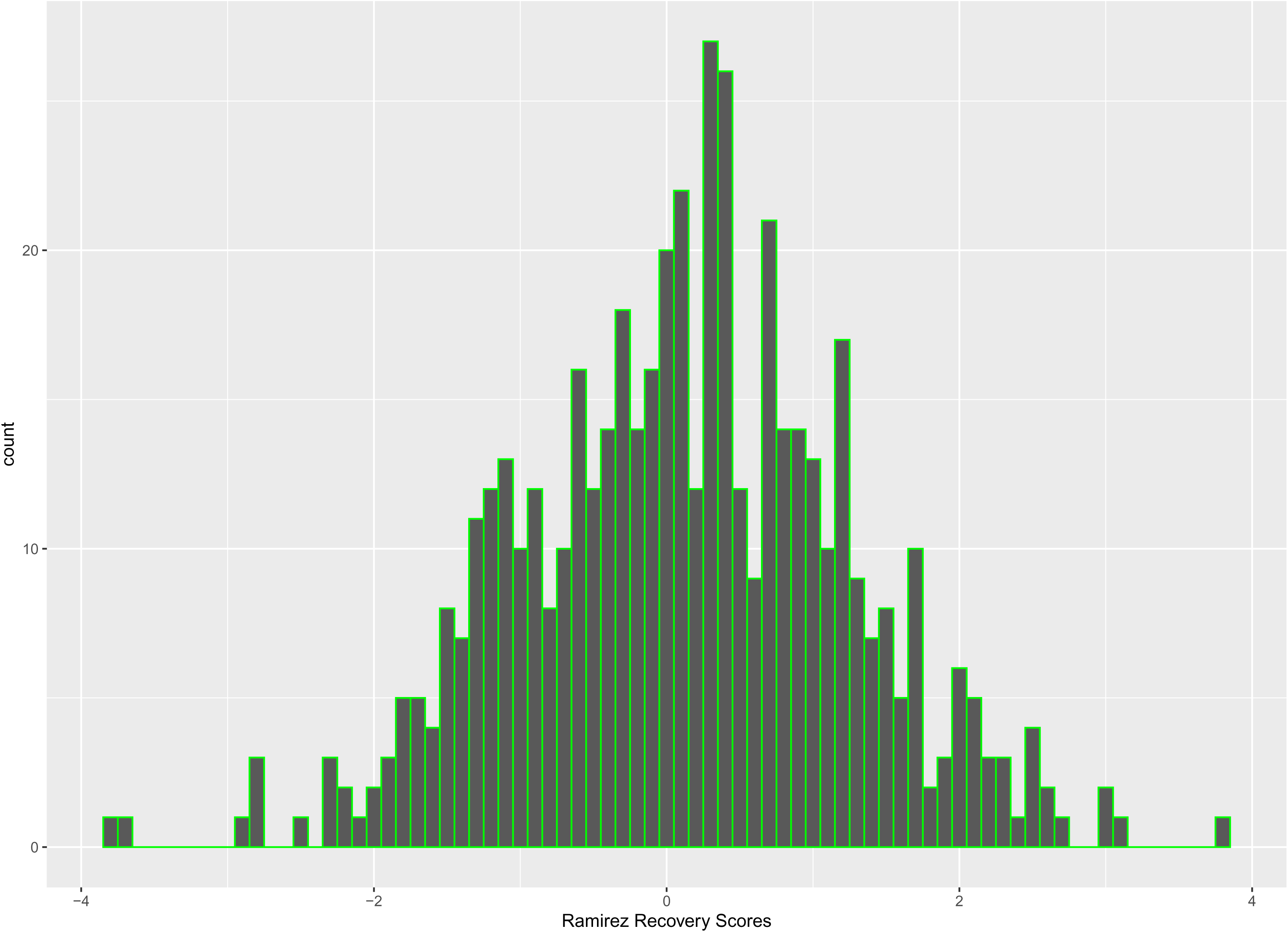
Distribution of the polygenic scores from the two studies in the European super-population of the 1000 genomes

**Supplementary Figure 4A:**
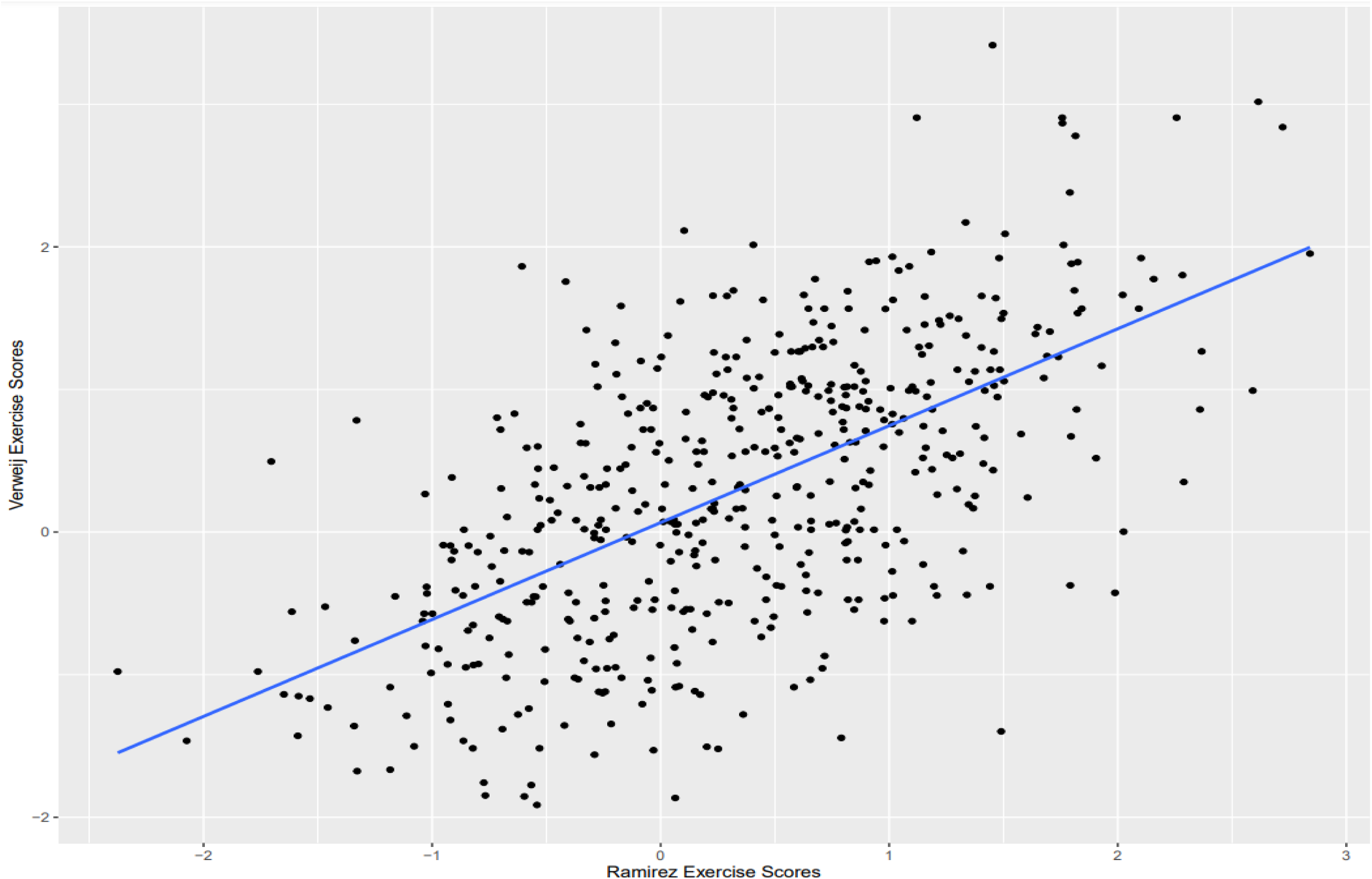
Scatterplots of exercise scores from the two studies in the 1000 genomes European superpopulation

**Supplementary Figure 4B-4F:**
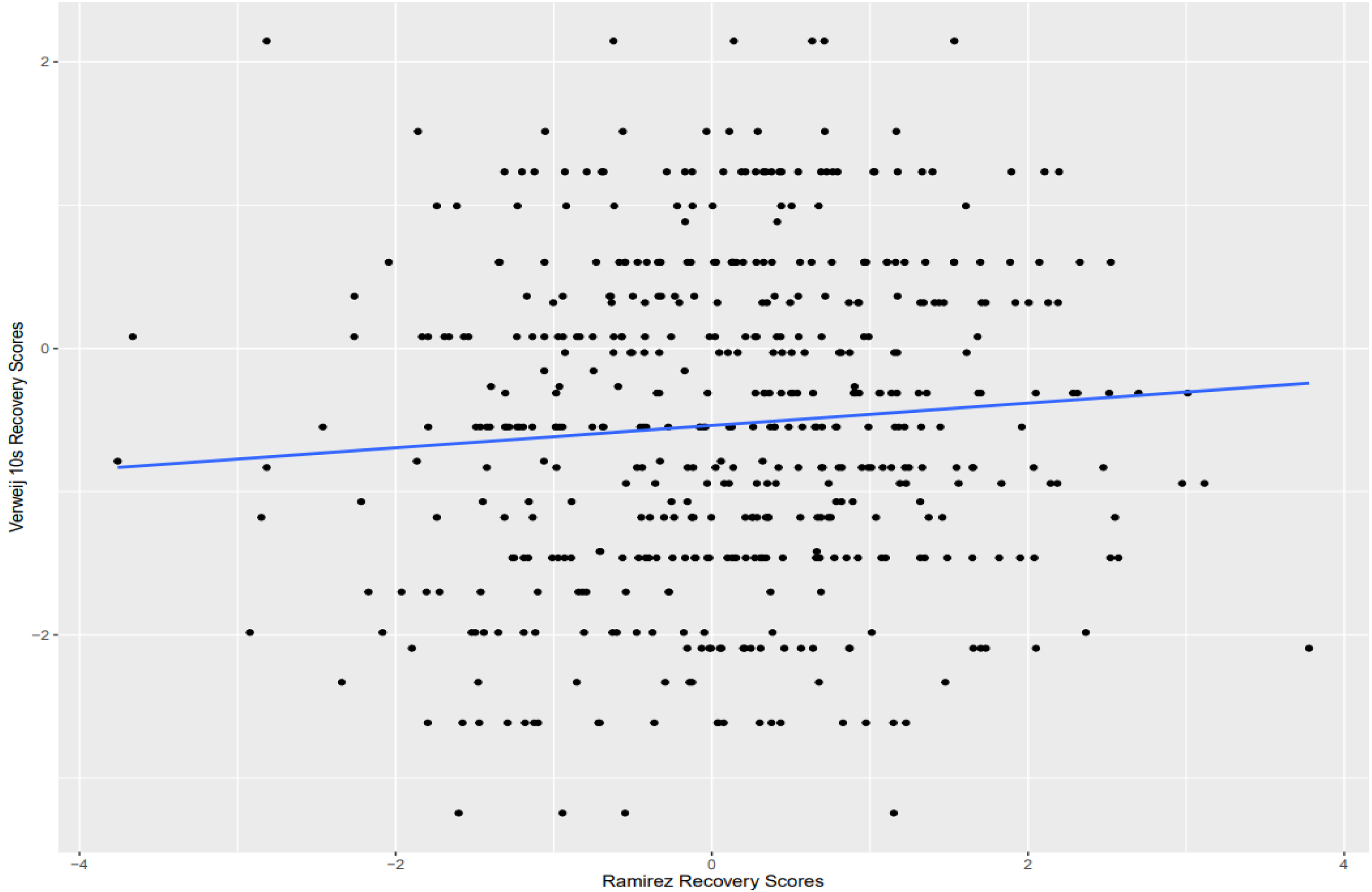
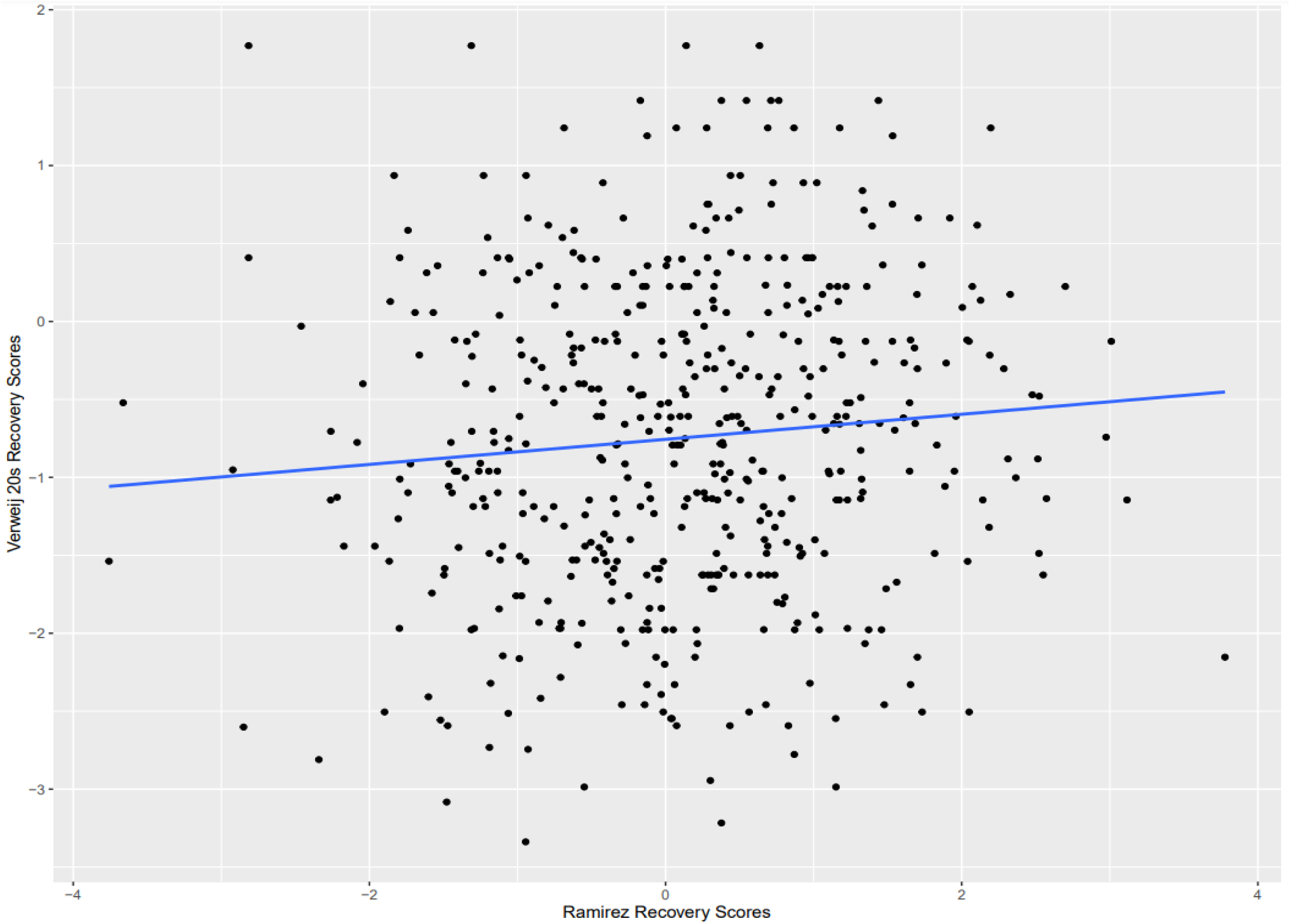
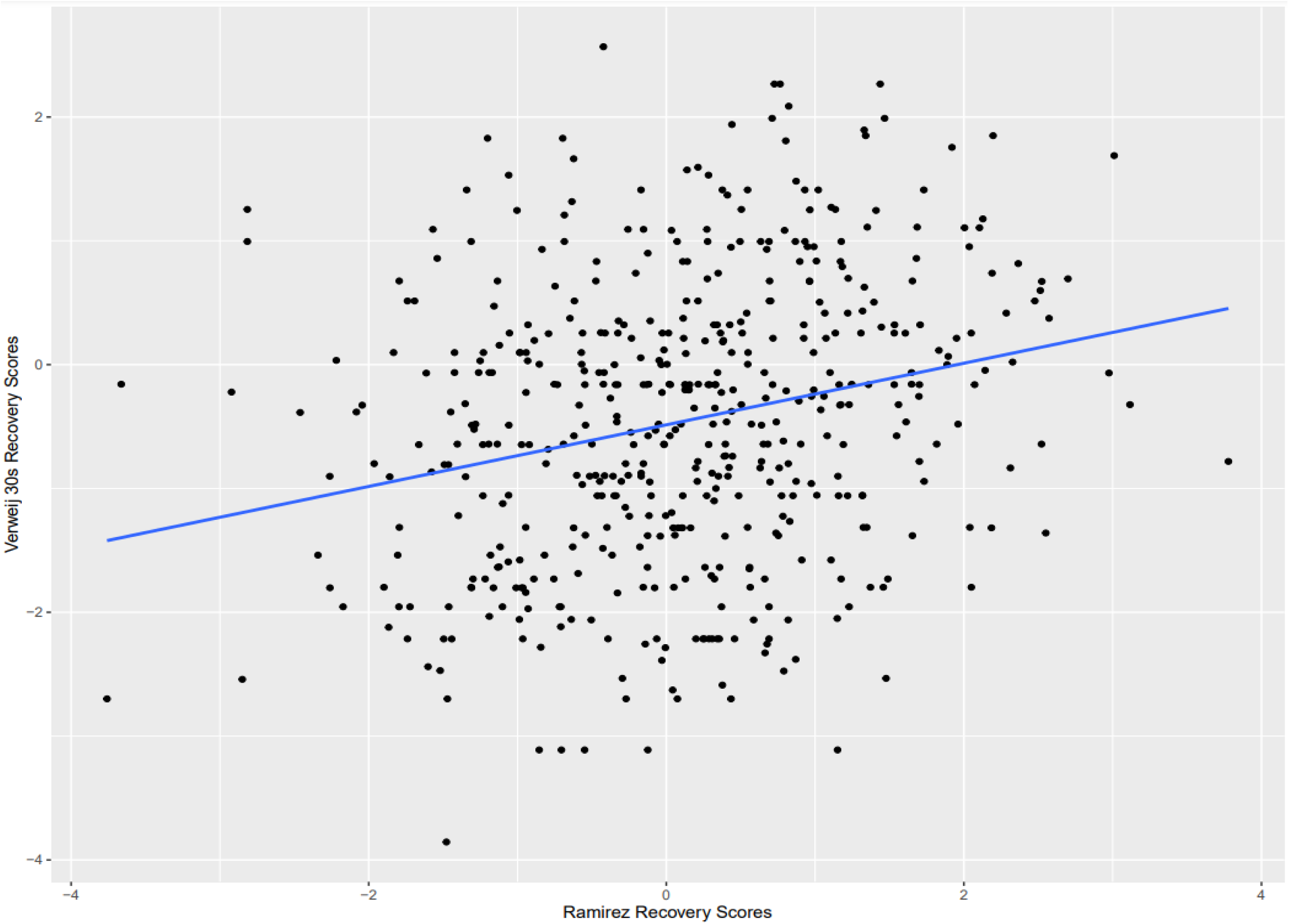
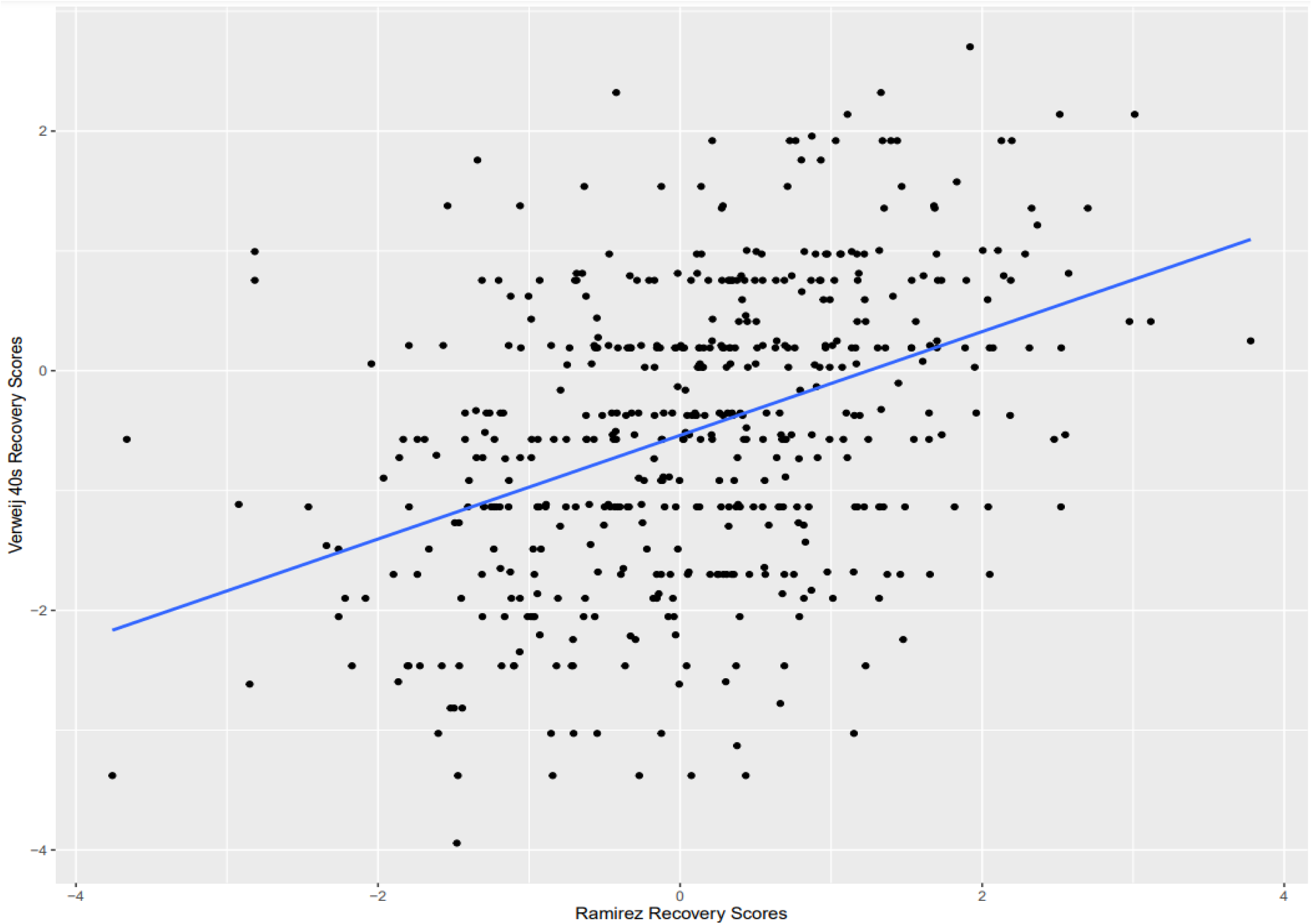
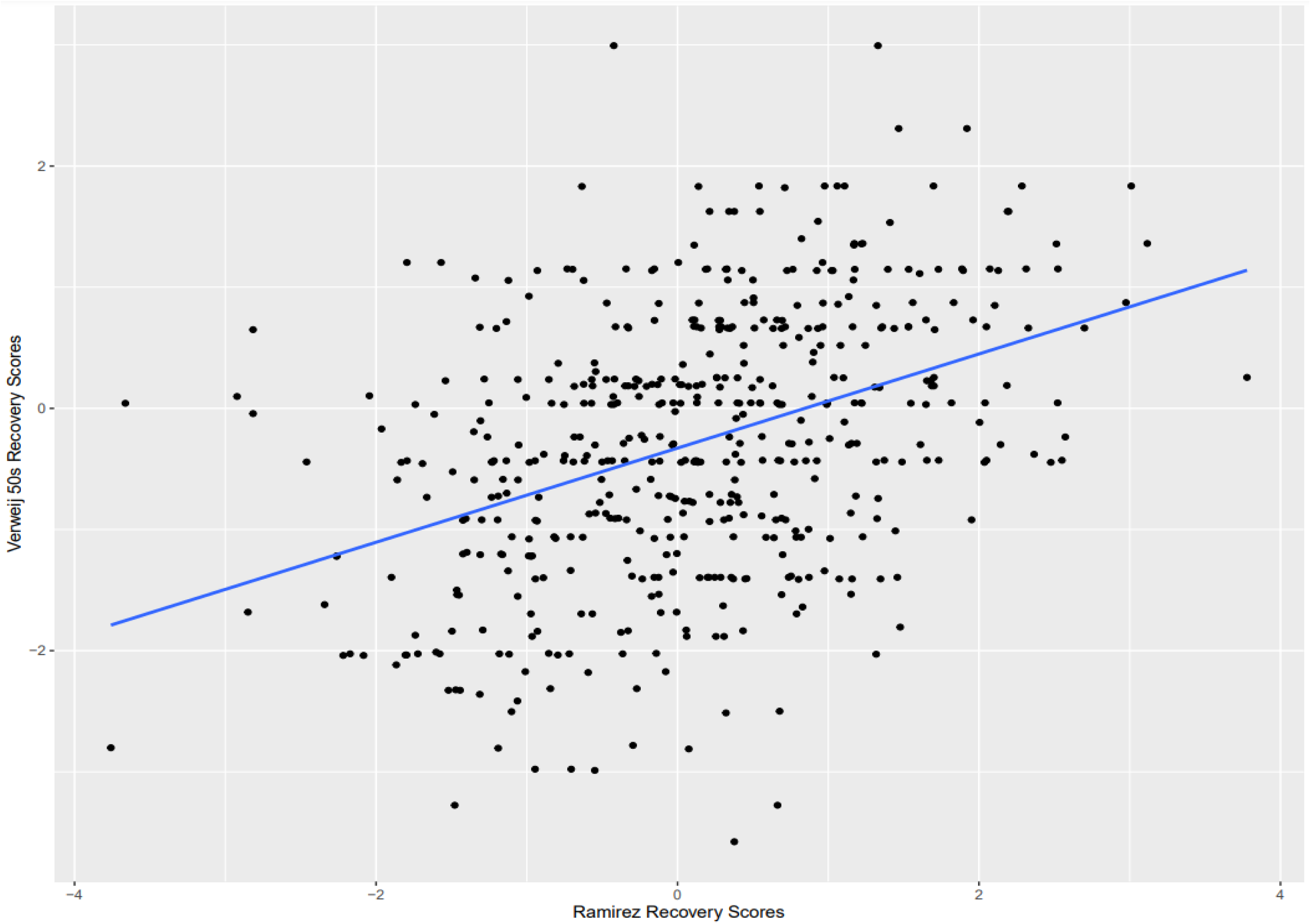
Scatterplots of recovery scores from Ramirez vs the 5 recovery scores from Verweij study in the 1000 genomes European superpopulation

**Supplementary Figure 5A:**
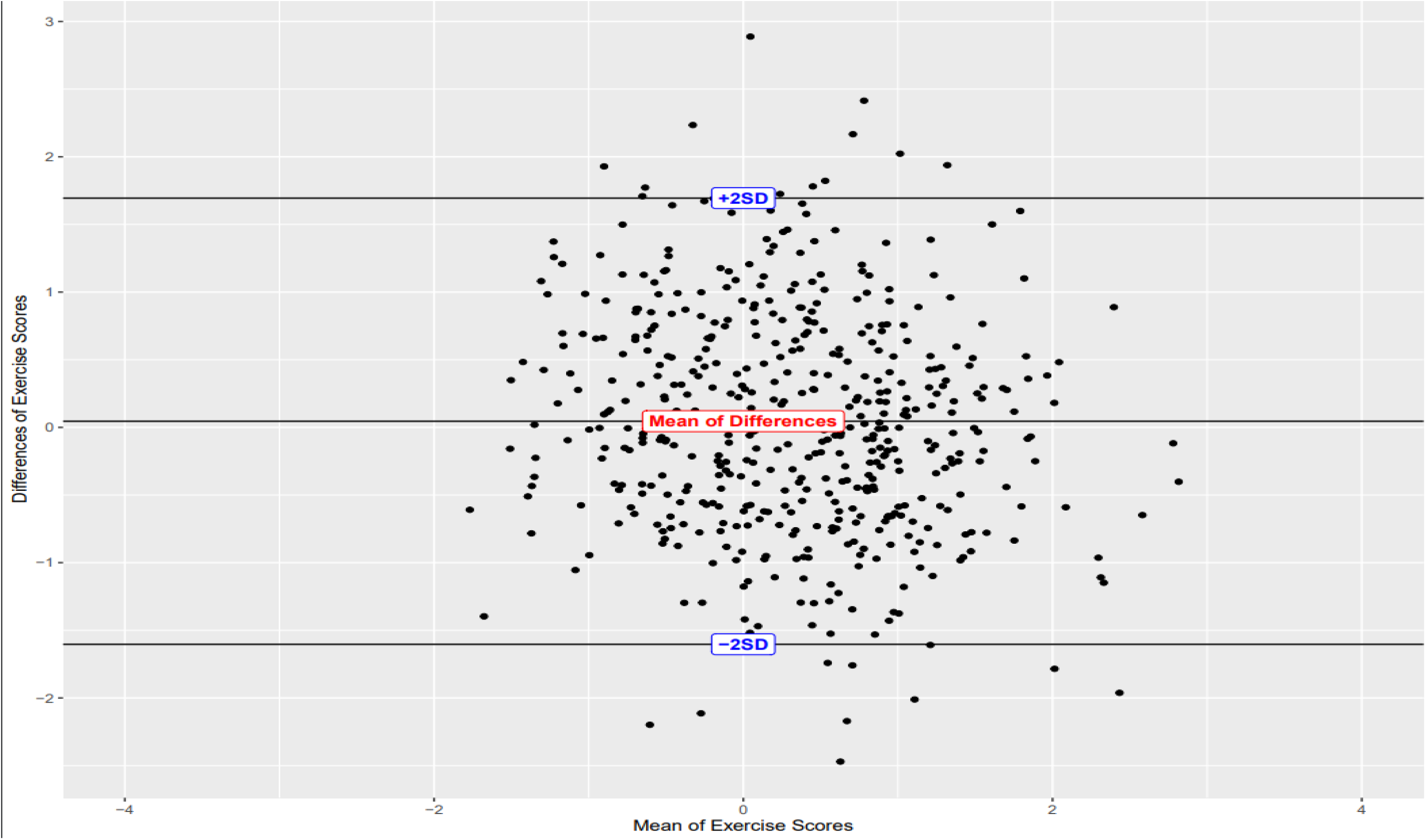
Bland Altman Plots of Exercise Scores of the two studies. (The differences are Ramirez scores-Verweij Scores). Scores were standardized to means of 0 and SD of 1.

**Supplementary Figure 5B-5F:**
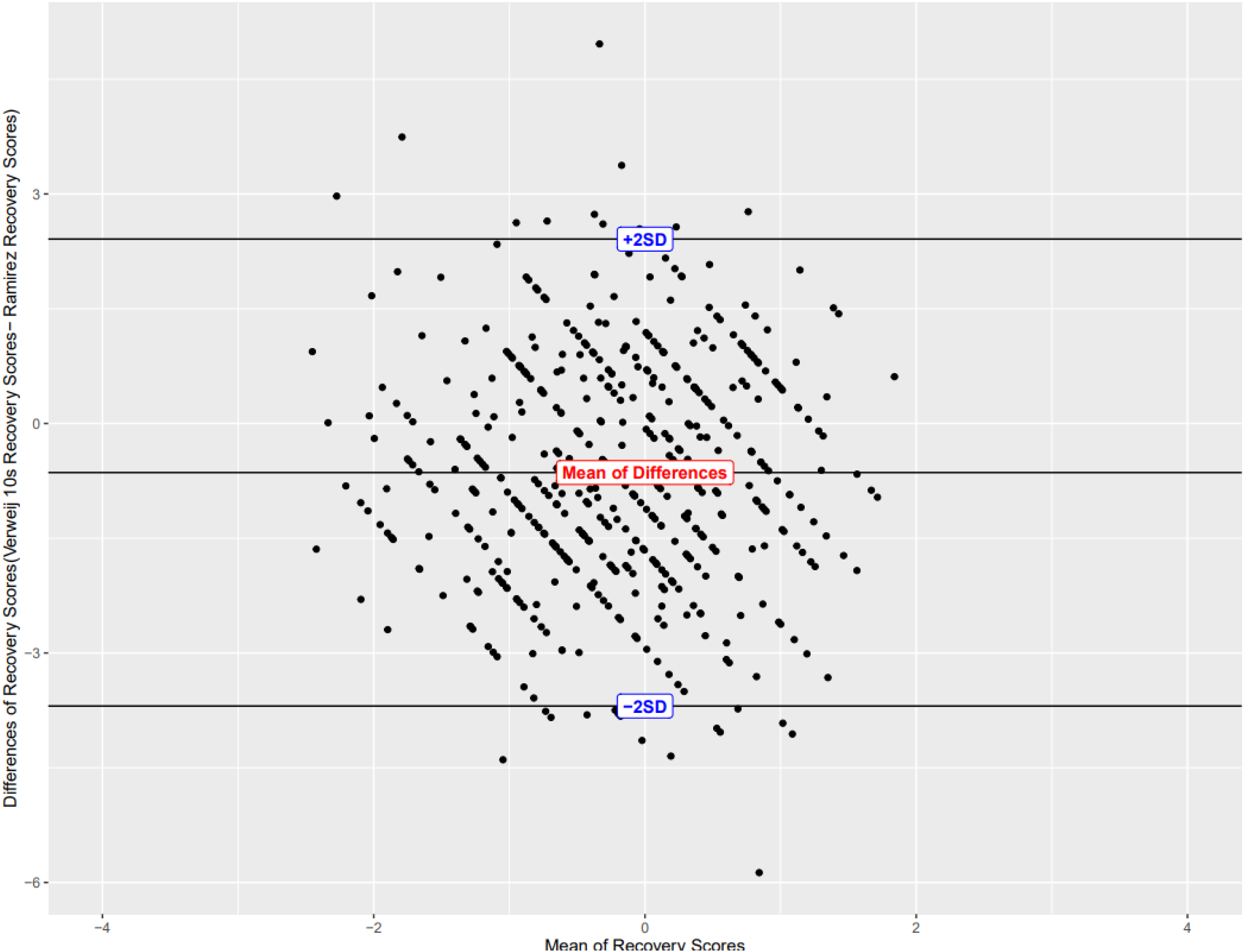
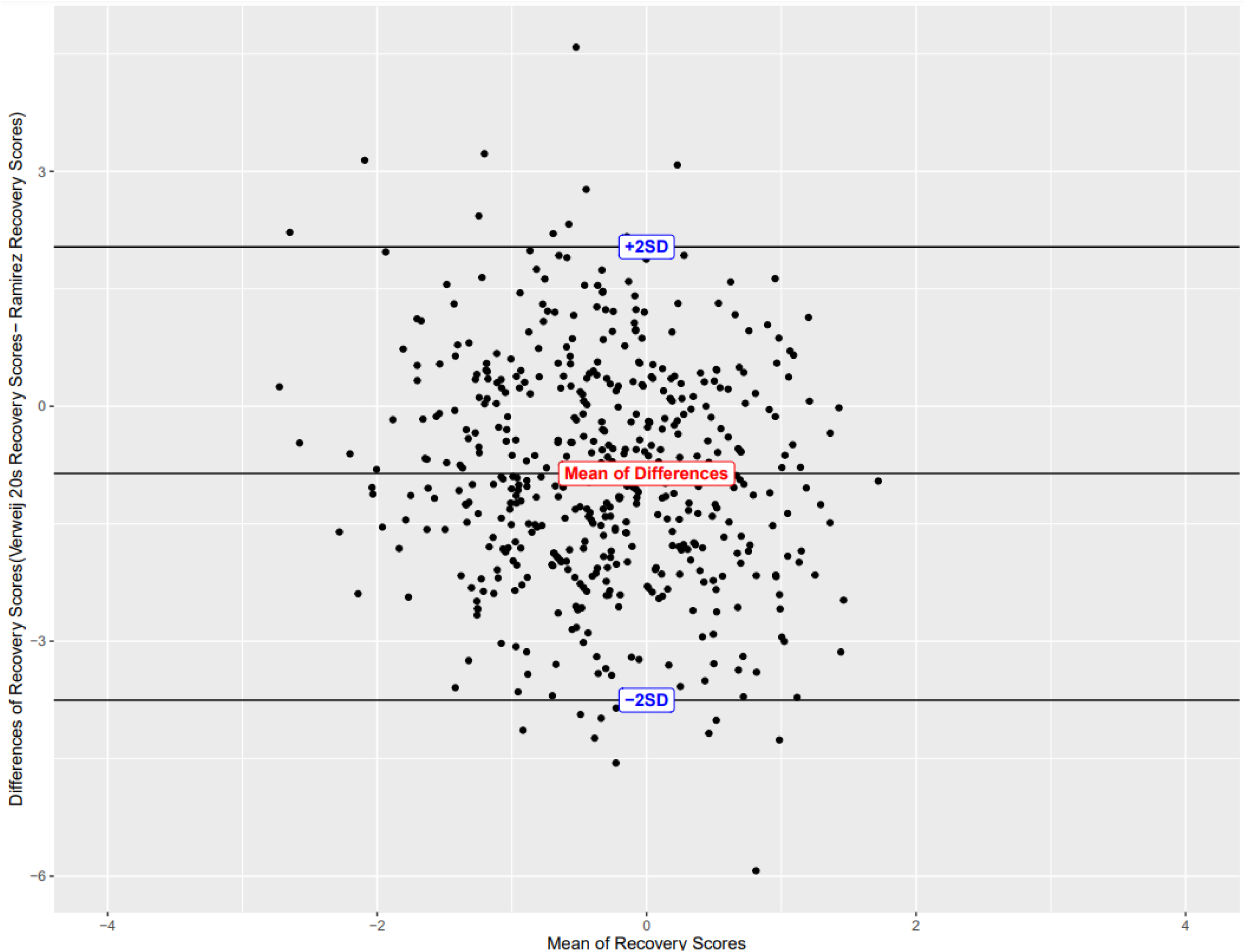
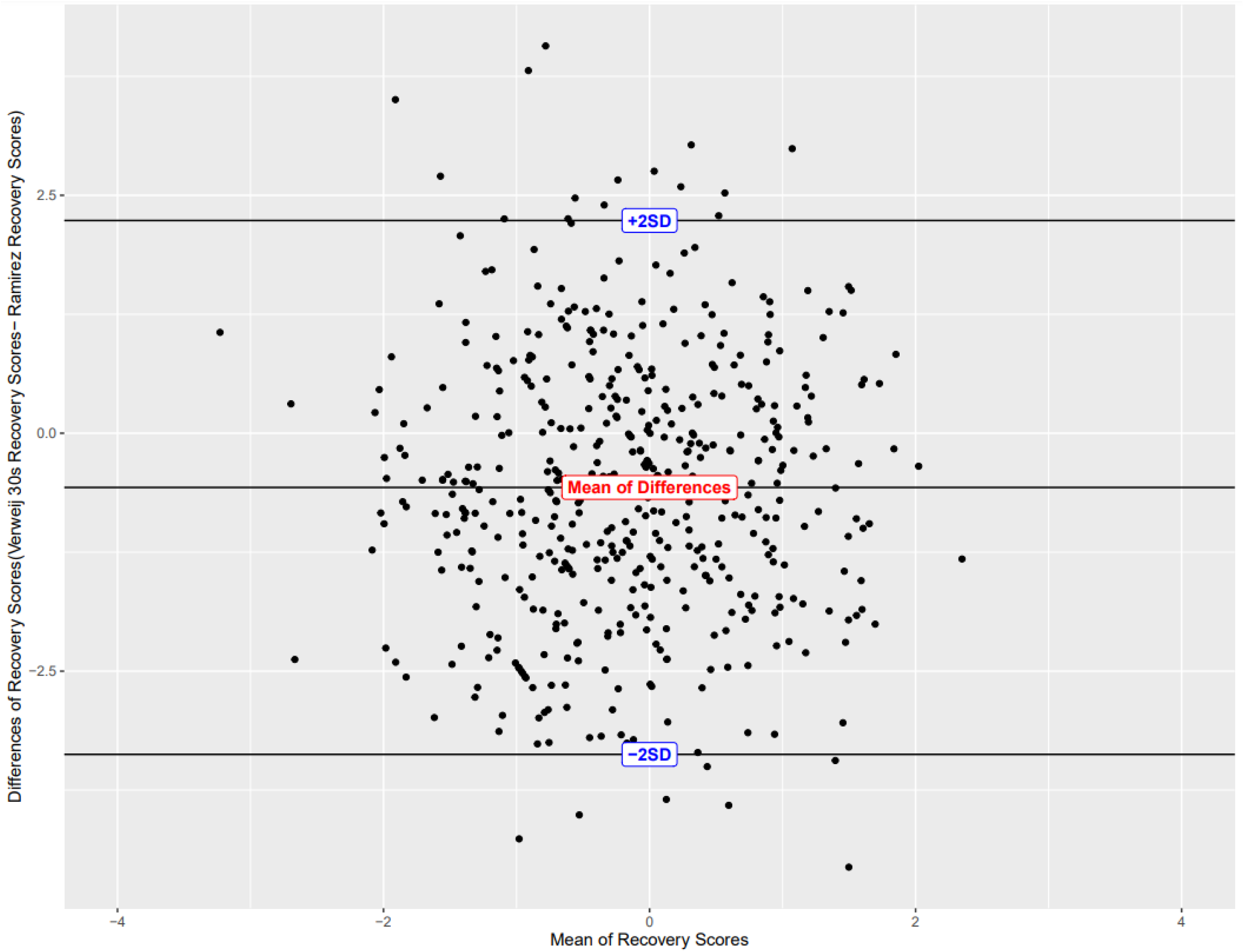
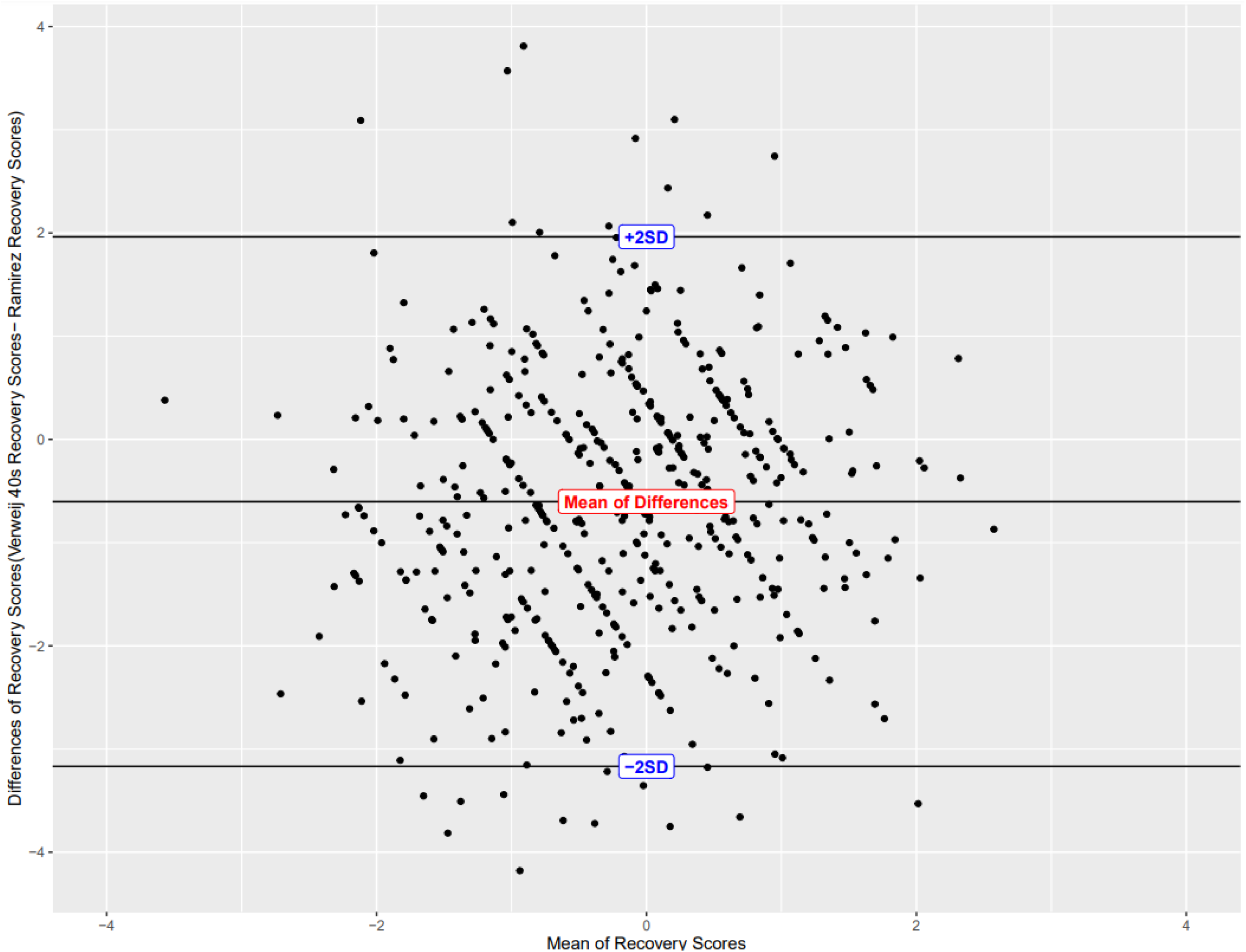
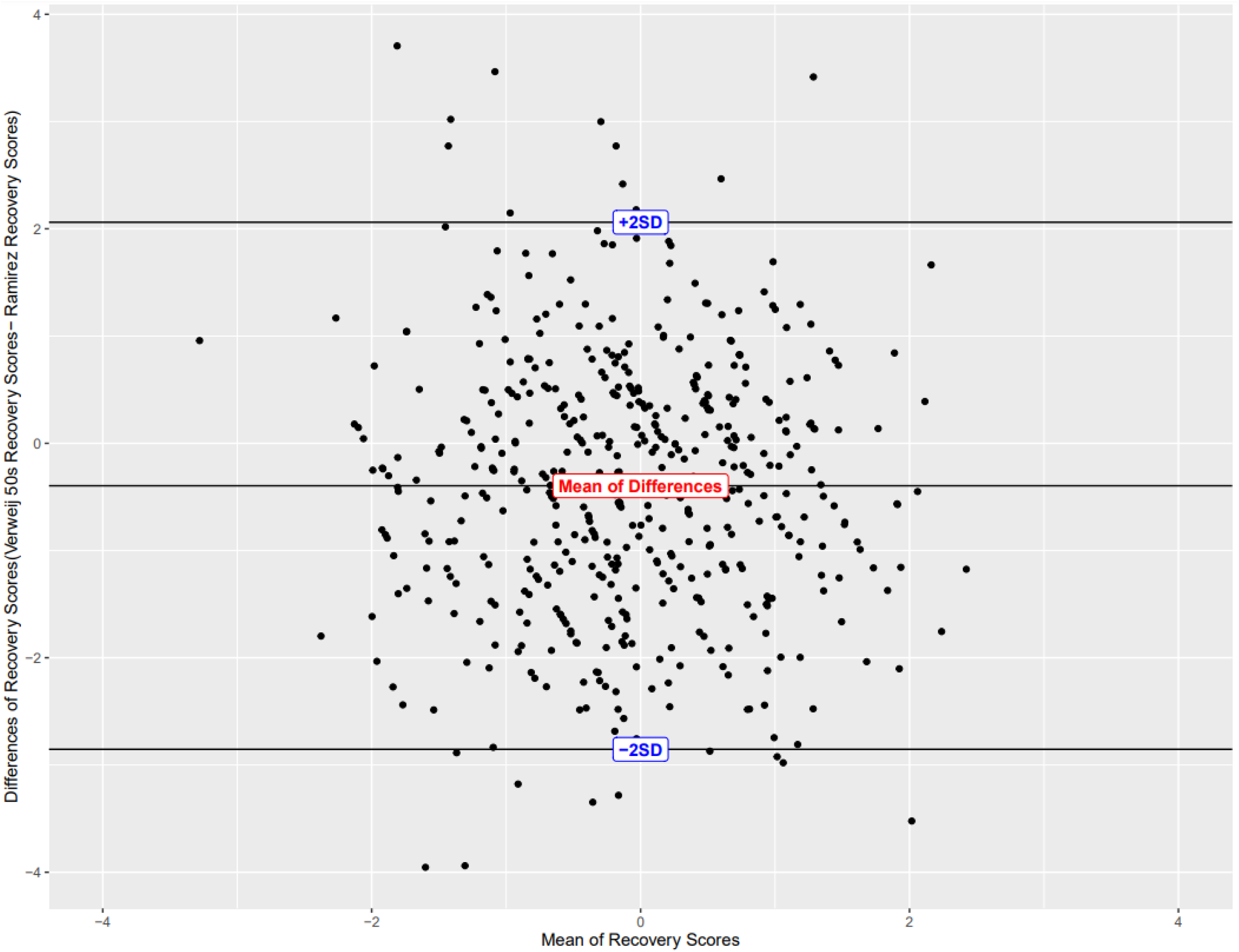
Bland Altman Plots of Recovery Scores of the Ramirez study compared with the 5 different recovery scores from Verweij study. (The differences are Ramirez scores-Verweij Scores). Scores were standardized to means of 0 and SD of 1.

## Appendix A Search Strategy for OVID Medline

1. genetics/ or exp genetic research/ or exp genetics, population/ or exp genomics/ or exp human genetics/
2. genetics.tw,kf.
3. Genome-wide association study.tw,kf.
4. Genome-wide association scan.tw,kf.
5. GWAS.tw,kf.
6. WGAS.tw,kf.
7. GWA study.tw,kf.
8. WGA study.tw,kf.
9. WGA scan.tw,kf.
10. GWA scan.tw,kf.
11. 1 or 2 or 3 or 4 or 5 or 6 or 7 or 8 or 9 or 10
12. Heart Rate/
13. RR interval.tw,kf.
14. heart rate.tw,kf.
15. 12 or 13 or 14
16. exp Exercise/
17. exercise.tw,kf.
18. (physical adj3 activity).tw,kf. 19. 16 or 17 or 18
19. 11 and 15 and 19

TW, KF refer to title and keyword fields.

